# Placental molecular subtypes of severe preeclampsia reveal divergent aging trajectories and fetal growth outcomes

**DOI:** 10.64898/2026.06.02.26354756

**Authors:** Yuheng Du, Paula A Benny, Shayanki Lahiri, Fadhl M. AlAkwaa, Qianhui Huang, Yuansen Liu, Cameron B Lassiter, Joshua Astern, Jonathan Riel, Lana X Garmire

**Author notes:** Department of Biomedical Informatics and Data Science, University of Alabama Birmingham, Birmingham, AL, USA.

## Abstract

Severe preeclampsia (sPE) is a major cause of maternal and fetal morbidity worldwide, yet its placental molecular heterogeneity remains poorly defined by current clinical diagnosis. To resolve the molecular architecture of sPE, here we integrated DNA methylation and proteomic profiling from a multi-ethnic cohort of 444 placentas from the Hawaii Biorepository (HiBR), including 169 sPE cases, matched preterm controls and full-term controls. To address cellular heterogeneity in bulk placental tissue, we developed HOMED (Hierarchically Optimized Methylation Deconvolution), a single-cell-guided hierarchical framework for inferring placental cell-type composition from DNA methylation data. HOMED-adjusted integrative analyses identified extensive subtype-specific alterations involving hypoxia, angiogenesis, immune activation, trophoblast differentiation and metabolic remodeling. Molecular stratification revealed two reproducible sPE subtypes with divergent placental aging trajectories. One subtype exhibited a pre-mature placental state marked by accelerated placental aging, whereas the other displayed slower accelerated placental aging but a substantially increased risk of small-for-gestational-age birth (P = 0.028). These subtypes were independently replicated across six external cohorts and further supported by proteomic signatures achieving a classification accuracy of 0.88. Integrative epigenomic and proteomic analyses linked the growth-restricted subtype to hypoxia-associated glycolytic remodeling, suggesting distinct pathogenic mechanisms underlying clinically diagnosed sPE. Together, our findings redefine severe preeclampsia as a biologically heterogeneous placental disorder composed of molecularly distinct subtypes with divergent aging trajectories and fetal growth outcomes, providing a framework for mechanism-based stratification and precision obstetric medicine.

## Introduction

Preeclampsia affects up to 8% of pregnancies and remains a leading cause of maternal and perinatal death^1^, contributing to an estimated 76,000 maternal and 500,000 neonatal deaths each year^2^. Severe preeclampsia (sPE), defined by severe-range hypertension with maternal end-organ dysfunction or uteroplacental compromise, represents the high-risk end of this spectrum^3,4^. Yet, sPE is currently managed as a monolithic clinical entity, diagnosed by convergent maternal symptoms rather than by specific underlying placental pathologies. Consequently, the molecular heterogeneity within clinically defined sPE, and its relationship to fetal growth restriction, placental maturation, and disease pathogenesis, remains poorly resolved.

Placenta is a vital temporary organ connecting the fetus to the mother’s uterus during pregnancy. It protects the baby through the maternal-fetal interface for nutrient exchange, endocrine signaling, and immune tolerance^5^. The placenta supports fetal growth through chorionic villi, branching structures bathed in maternal blood within the intervillous space. Each villus is composed of trophoblasts in the outer layer and the stromal core inside, including fetal endothelial cells, stromal fibroblasts, placenta-resident macrophages known as Hofbauer cells^6^. Among trophoblasts, cytotrophoblasts (CTBs) act as progenitors, which either fuse to form multinucleated syncytiotrophoblasts (SCTs) on exchange surface of the villous placenta, or differentiate into extravillous trophoblasts (EVTs) to invade the maternal decidua and remodel spiral arteries for circulation^5^. Defective trophoblast invasion, impaired spiral artery remodeling, hypoxia, and angiogenic imbalance are known features of PE^7^. However, whether these processes represent a single disease continuum or distinct disease programs within sPE remains unclear.

Epigenome-wide DNA methylation is well suited to study pregnancy complications including PE, owing to its tissue specificity, long-term stability, and capacity to capture both developmental programming and pathological insults^8–10^. However, bulk-level methylation study in placenta is analytically extraordinarily challenging due to multiple issues. First, complex tissues such as placenta are intrinsically vulnerable to confounding due to the heterogeneity in cell type proportions among individuals^11–14^, which are further significantly affected by diseases such as sPE but ignored by many studies unfortunately^15,16^. More importantly, placenta is a dynamic organ where the cell type composition changes significantly and continuously along the gestational age, which is also often overlooked by studies^17^. Without proper statistical adjustment to the cell-type variations and gestational age, the conclusions from epigenome-wide association of placentas may biasedly reflect placental compositional remodeling rather than the intended disease-associated epigenetic changes^18–20^. Moreover, methylation records regulatory and developmental memory rather than the direct biological functional readout. Proteomics provides complementary functional output that executes placental physiology and pathology^21^. Integration of these two omics can reveal much more complete biological programs that shape placental dysfunction and fetal outcome.

To identify robust molecular subtypes and cellular and molecular heterogeneity that may underlie sPE, we herein integrated placental DNA methylation and proteomic profiles from 444 placentas in the Hawaii Biorepository, including 169 sPE cases, 106 matched preterm controls and 169 full-term controls. To directly address the above mentioned analytical challenges in large-scale epigenome-wide association study, particularly in placentas, we developed HOMED (Hierarchically Optimized Methylation Deconvolution), a single-cell-guided hierarchical framework for inferring cell-type composition from DNA methylation data. Consequently, we identified two reproducible sPE molecular subtypes with distinct cell compositions, placental aging acceleration trajectories and fetal growth risks. These subtypes were validated by external cohorts as well as proteomics measurement. Moreover, integrative epigenomic and proteomic analyses further linked the growth-restricted subtype to hypoxia-associated glycolytic remodeling and thrombo-inflammatory response, confirming distinct pathogenic mechanisms contributing the phenotypes. By identifying distinct subtypes that manifest clinically similar sPE symptoms, this work provides a basis for developing subtype-specific biomarkers and moving toward precision obstetric medicine.

## Results

### Overview of severe preeclampsia placenta cohort from Hawaii Biorepository (HiBR)

HiBR is a unique population-based multi-ethnic in Hawaii, with over 9,000 mother-child pair specimen collections. To investigate the heterogeneous molecular landscape of sPE, we conducted a nested multi-omics case and control study on 444 placentas. This cohort includes 169 sPE cases, 106 preterm controls with matched propensity-scores on critical clinical factors including gestational age, maternal age, pre-pregnancy BMI, parity, ethnicity and smoking, as well as 169 full-term controls that matched on the same variables as in preterm controls except the gestational age (**Figure 1; Supplementary Table 1**). The preterm controls are designed specifically to remove gestational age as a major confounder to study sPE, where preterm delivery is often the clinical decision made to mitigate maternal risks of sPE^22^. As confirmation of the study design, gestational age, most baseline demographic variables including maternal age, ethnicity, smoking status and obstetric variables including gestational age, parity, abruption, baby sex, are balanced between cases and controls (**Figure 2a,b,d,e,h-j,l**). Other variables, including maternal pre-pregnancy BMI, chronic hypertension, gestational diabetes and small-for-gestational-age birth, are significantly higher in sPE as expected (**Figure 2f,g,k**), demonstrating the maternal and fetal growth burden of sPE^23–25^.

**Figure 1.**
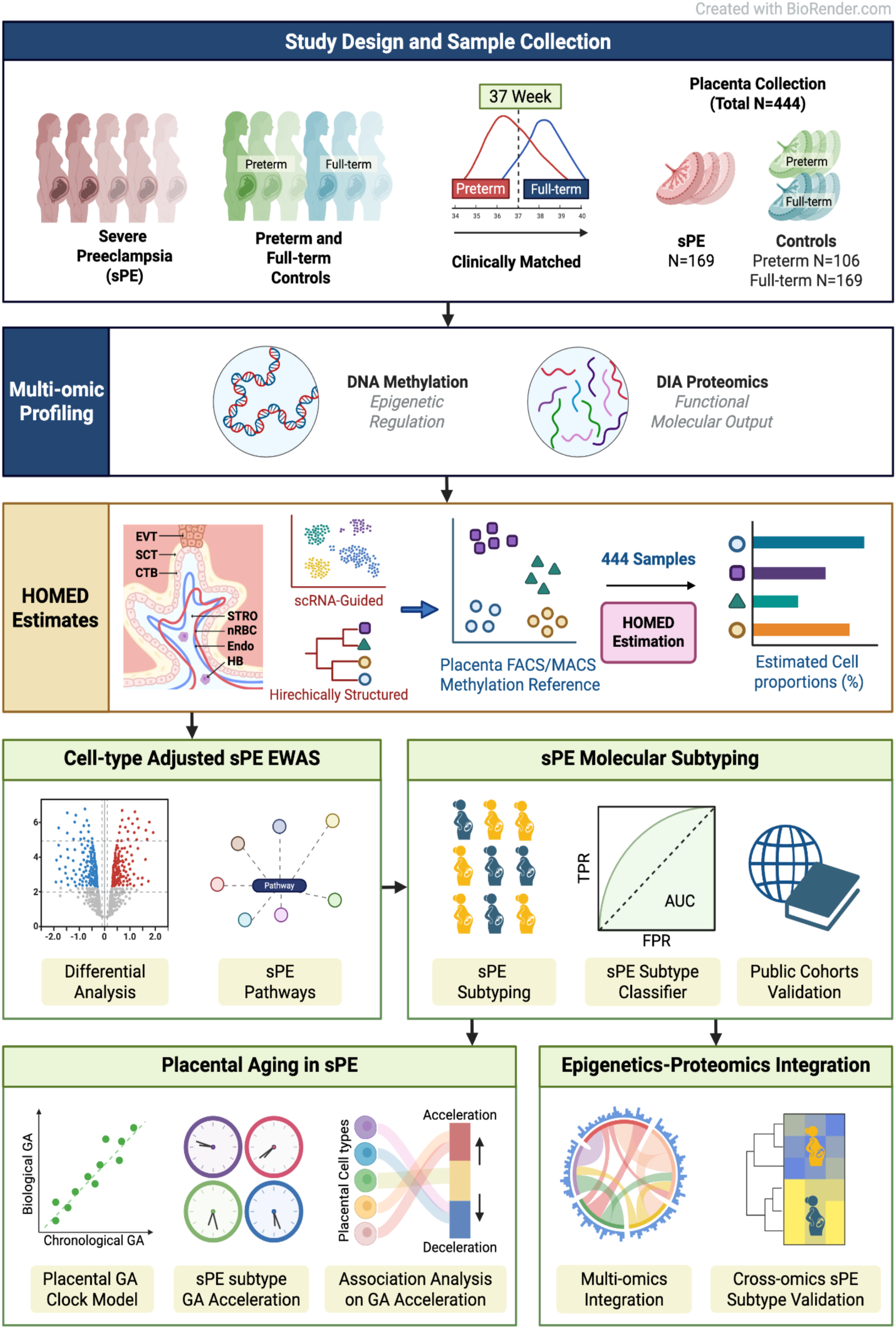
Overview of study design and multi-omic profiling workflow. Placental tissues were collected from 444 pregnancies in HiBR, including those from severe preeclampsia (sPE, n=169), propensity-score matched pre-term deliveries (n=106), and full-term deliveries (n=169). DNA methylation profiling was performed using the Illumina Infinium EPIC array, and proteomic profiling from a subset of the same samples (n = 158, including 78 sPE and 80 GA-matched controls) was performed by Data-Independent Acquisition (DIA) mass spectrometry. To decipher placental cell-type heterogeneity associated with sPE, we developed HOMED (Hierarchically Optimized Methylation Deconvolution), an scRNA-Seq-guided, hierarchical relationship-based bulk DNA methylation deconvolution method. The inferred cell-type information was applied to differential methylation (DM) analysis, pathway analysis, and identifying epigenetic subtypes of sPE. Then the placental GA clock model was constructed and applied to sPE subtypes. The differences in predicted GA between the sPE subtypes were associated with specific cell types in the placenta, providing mechanistic insights. Lastly, DNA methylation and proteomics integration was performed, and the sPE epigenetic subtypes were validated by additional DNA methylation public datasets, and by the proteomics data.

**Figure 2.**
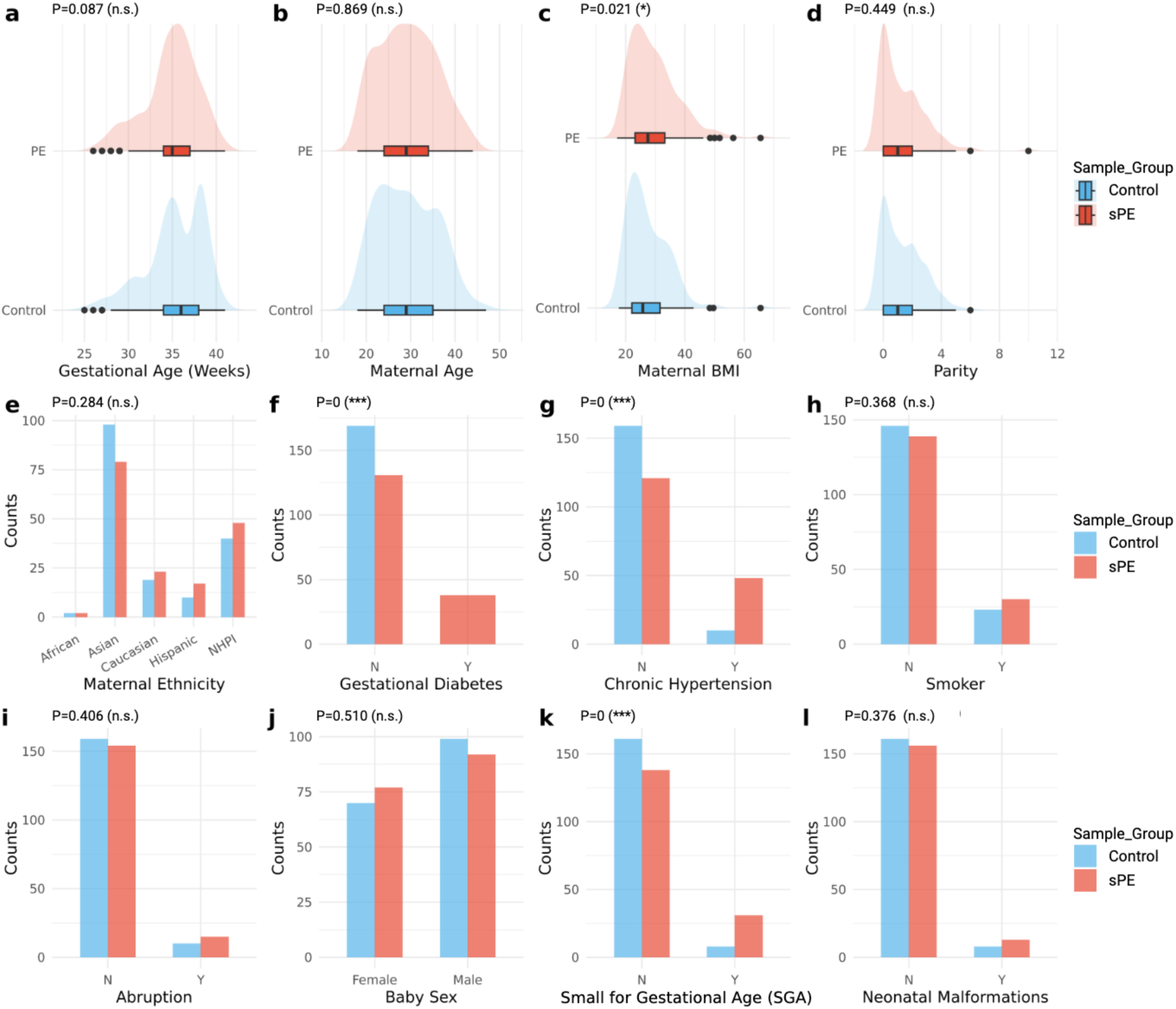
HiBR placenta cohort demographic characteristics. Pre-term controls are matched to sPE cases by propensity scores. (a-d) Distributions of gestational age (GA) in weeks (a), maternal age (b), maternal body mass index (BMI) (c) and parity (d) across GA-matched controls and sPE samples. (e) Ethnicity composition across study groups. (f-l) Comparison of discrete clinical variables between the GA-matched controls and sPE samples, including (f) gestational diabetes prevalence, (g) chronic hypertension, (h) smoking status, (i) placental abruption, (j) fetal sex distribution. (k) newborn small-for-gestational-age (SGA) status, and (l) neonatal malformation.

For each patient in the HiBR placenta cohort, we retrieved placental tissues from the standardized mid-villous parenchymal region. We generated genome-wide DNA methylation profiles for all 444 placentas using the Illumina EPIC array, followed by sex verification, probe filtering, normalization, batch correction and removal of cross-hybridizing probes (**Supplementary Figure 1a-c**). To link epigenetic variation with functional molecular output, we further performed DIA-based proteomics on 158 propensity-score-matched placentas, including 78 sPE cases and 80 controls, with preprocessing and quality control summarized in **Supplementary Figure 1d**.

### Cell-type deconvolution reveals trophoblast and villous core abnormalities in severe preeclampsia

Methylation profile in bulk tissues such as placenta affected by sPE is an aggregated measurement of methylation from all cell types within, reflecting both disease-associated methylation changes and cell-type composition change. Thus removing the effect due to cell-type variations in epigenome-wide association study of sPE is critical. Cohort scale cell-type proportion estimation is conventionally done by deconvolution with cell-type specific DNA methylation reference^26,27^. However, deconvolution is especially challenging in the placental tissue, where the principal trophoblast lineages - cytotrophoblasts (CTBs), syncytiotrophoblasts (SCTs) and extravillous trophoblasts (EVTs) - share closely overlapping methylation profiles. As a result, it has been very difficult for conventional reference-based methods to accurately estimate the proportions of cell types from trophoblasts^28^ and placentas in general.

To overcome this major challenge, we developed HOMED, a single-cell RNA-seq guided methylation deconvolution framework that explicitly considers the cell differentiation lineage (**Figure 3a; Methods**). In placenta, HOMED uses purified methylation data from seven major cell types as the input: CTB, SCT, EVT, stromal cells, endothelial cells, Hofbauer cells and nucleated red blood cells (nRBCs) (**Figure 3b; Supplementary Figure 2; Supplementary Table 2**). Worthy noticing, we also expanded the cell types to include other immune cell types in blood with single cell RNA-seq data, but found the inferred proportions of these cell types are neglectable^29^ (**Supplementary Figure 3**). Next, HOMED performs deconvolution in hierarchical structured stages. It first conducts coarse-level deconvolution by combining CTB, SCT and EVT into one general trophoblast group, and estimating it along other four major placental cell types. In the second stage, it further decomposes trophoblast into CTB, SCT and EVT for iteratively refined cell proportion estimation. At each refinement step, methylation-based cell proportion estimates are constrained by single-cell RNA-seq reference (**Figure 3c**) guided cell proportion estimation on the RNA-seq data modality from the placenta samples with coupled RNA-seq and DNA methylation data (**Figure 3d)**.

**Figure 3.**
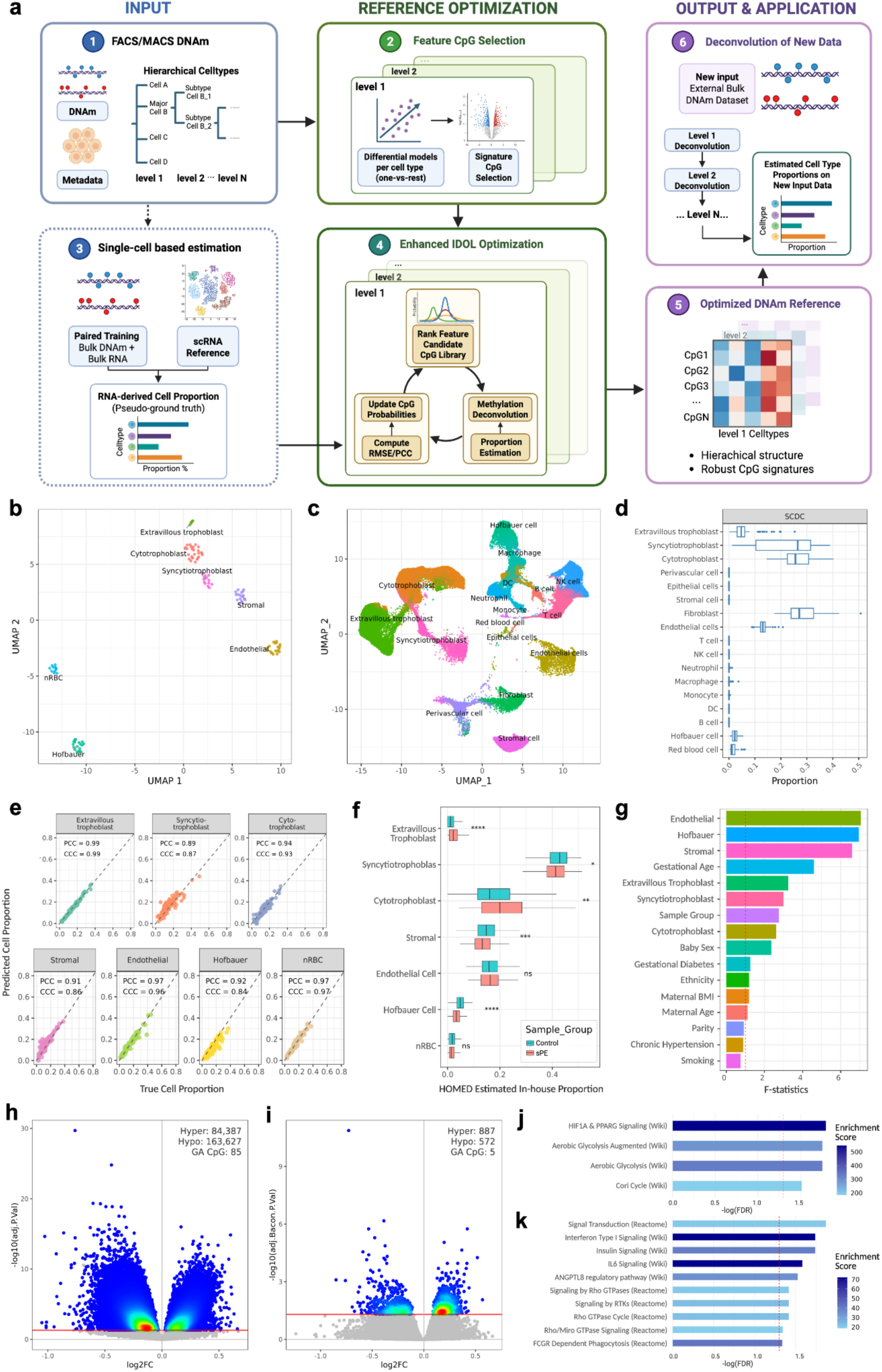
Placental DNA methylation deconvolution by the newly developed HOMED computational method. (a) The Schematic overview of HOMED (Hierarchically Optimized Methylation Deconvolution). It applies the FACS/MACS-sorted cell-type specific methylation profiles to construct hierarchical CpG signature matrices for methylation-based cell-type deconvolution. HOMED highlights using single-cell RNA-seq data-based references to guide constraining the cell-type proportion estimation of bulk DNA methylation data in the paired bulk DNA methylation-transcriptomic datasets. Step 1: Integrate FACS/MACS-sorted DNAm profiles to construct the initial methylation reference matrix. Step 2: Identify initial cell-type-specific CpG methylation marker candidates using differential methylation analysis, where the cell type of interest is compared with all the rest cell types (one-verus-rest). Step 3: Estimate cell proportions from paired bulk DNAm-RNA datasets, using single-cell RNA-Seq atlas data based deconvolution methods (e.g. SCDC) on the bulk RNA data. Step 4: Iteratively optimize CpG cell-type specific markers by minimizing RMSE and maximizing PCC against RNA-derived cell proportion estimates in Step 3, until convergence. Step 5: Generate the optimized hierarchical DNA methylation reference matrix. Step 6: Apply the optimized DNA methylation reference matrix to deconvolute new bulk methylation datasets. (b) Visualization of cell-type-specific methylation reference from the placenta in step 1, by UMAP plot from integrated FACS/MACS-sorted cell-type data from previous studies (Hu 2015; Lucas 2019; Yuan 2021; Campbell 2024). (c) UMAP visualization of the integrated placental single-cell RNA-seq atlas in Step 3, from six public studies (Campbell 2023; Li 2022; Han 2020; Liu 2018; Vento-Tormo 2018; Cao 2020). (d) Placenta cell proportions from studies GSE98224 and GSE73377, estimated by SCDC using single-cell RNA-seq atlas data. Seven major non-zero cell types are identified. (e) Performance evaluation of the HOMED method using simulation experiments. Scatter plots show predicted (y-axis) versus simulated ground-truth (x-axis) cell proportions of placentas. Metrics of pearson correlation coefficient (PCC) and the concordance correlation coefficient (CCC) values are shown. (f) HOMED-estimated cell proportions in our HiBR placenta cohort comparing the GA-matched controls and sPE placentas. (g) Source-of-variation analysis showing the relative contributions to global methylation variance from the estimated placental cell types and clinical variables. (h) Differential methylation analysis without cell-type adjustment. (i) Differential methylation analysis after HOMED-based cell-type adjustment and Bacon correction. The number of differential methylation CpG probes is significantly reduced with such adjustments. (j) Pathway enrichment of hypomethylated CpGs in the promoter regions (TSS200 and TSS1500 regions), which may indicate upregulation of these pathways. (k) Pathway enrichment analysis of hypermethylated gene body CpGs, which may relate to the upregulation of these pathways.

We first tested HOMED by artificial placenta mixture. It accurately estimates the above mentioned major placental cell type proportions, with Pearson’s correlation coefficient (PCC) of 0.89-0.99 and concordance correlation coefficient (CCC) of 0.84-0.99 (**Figure 3e**). We also demonstrate the consistent superior accuracy of HOMED over other similar single cell rna-seq guided deconvolution methods such as EPISCORE^30^ and scDeconv^31^, in a variety of tissues including placenta^32^. We then applied HOMED to the 444 placentas, and detected marked cellular remodeling in sPE (**Figure 3f**). Compared with all combined full-term and preterm controls, sPE group shows significantly expanded CTB and EVT proportions, but much reduced SCT, stromal, and Hofbauer compartments (P < 0.001). This pattern indicates overall disrupted trophoblast maturation in sPE, by retaining less mature trophoblast state and reducing its differentiation to SCT^13,33^. With stromal and Hofbauer depletion, sPE appears to show an overall imbalanced villous environment, characterized by loss of supportive stromal-immune compartments ^34^. However, such sPE vs. control group-level differences may not imply a uniform cellular state across all sPE placentas, and the heterogeneous sPE epigenome landscape will be revealed subsequently.

### Cellular and gestational age correction significantly reduce inflation in sPE-associated methylation analysis

To investigate the contributors to variations in DNA methylation, we conducted a source of variance analysis (**Figure 3g**). Placental cell proportions rank among the strongest sources of methylation variation, supporting the need to account for adjusting cell type variations in placental epigenome-wide association study (EWAS)^11,19^ of sPE. Gestational age also contributes to more DNA methylation variation than sPE/control grouping, confirming the importance to adjust for gestational age in EWAS too. Consequently, we adjusted the EWAS model for placental cell compositions inferred by HOMED, gestational age and other clinical covariates. Residual test-statistic inflation was further corrected using BACON, an empirical-null framework for controlling bias and inflation in EWAS^35^ (**Supplementary Figure 4**). Such rigorous statistical adjustment drastically reduces genomic inflation by 170 folds (**Figure 3h-i),** highlighting the critical value of comprehensive correction for cellular and clinical confounders to uncover genuine sPE-associated methylation signals.

In total, we detected 1,459 sPE-associated CpGs, including 887 hypermethylated and 572 hypomethylated sites (**Supplementary Table 3**). Functional annotation of hypomethylated CpGs in the promoter regions shows enrichment for HIF1A signaling and aerobic glycolysis (**Figure 3j**), consistent with hypoxia-driven metabolic adaptation in sPE ^36^. Hypermethylated gene body sites are enriched with type I interferon signaling, IL-6 signaling, insulin signaling, Rho GTPase activity and Fcγ receptor-mediated phagocytosis (**Figure 3k**). As methylation in the gene body is positively related to gene expression^37^, this implicates immune activation and cytoskeletal remodeling at the maternal-fetal interface.

### Two sPE methylation subtypes are robustly identified and validated with divergent fetal growth outcome

To investigate the heterogeneity of placentas affected by sPE, we performed unsupervised clustering, after adjusting the methylation profiles by confounders including gestational age, gender, BMI, gestational diabetes, ethnicity, maternal age and smoking status. We observe two stable subtypes regardless of the clustering methods, and denote them as subtype 1 and subtype 2 (**Figure 4a; Supplementary Figure 5**). Although the subtypes are largely indistinguishable judging by conventional maternal and pregnancy-related clinical variables, they differ markedly in fetal growth outcome (**Supplementary Table 4**). Subtype 2 shows significantly elevated SGA risk (P < 0.05), with 25.3% of births classified as SGA compared with 11.6% in Subtype 1 and 4.9% in gestational-age-matched controls, corresponding to more than 2-fold and 5-fold increases, respectively (**Figure 4b**). In short, without epigome profiling these subtypes with divergent growth-restricted placental states would have been concealed by the same clinical sPE diagnosis.

**Figure 4.**
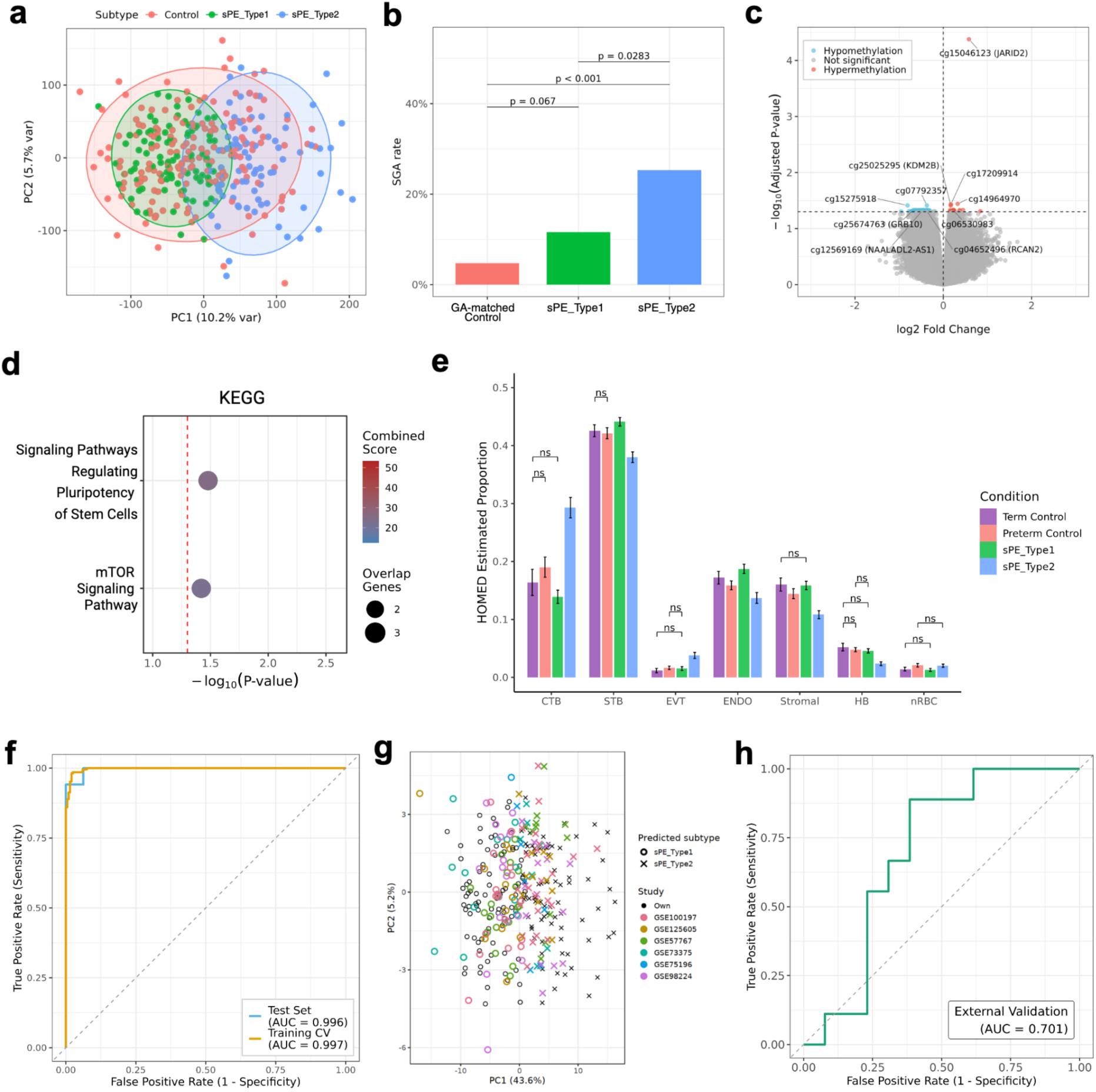
Identification of two epigenetic subtypes of placentas from sPE. (a) Principal component analysis (PCA) of confounder-adjusted methylation profiles showing two epigenetic sPE subtypes relative to controls. (b) Small-for-gestational-age (SGA) frequencies between the two sPE subtypes (sPE_Type1 and sPE_Type2), as well as from the GA-matched controls. P-values shown are calculated from Fisher’s exact tests. (c) Volcano plot of subtype-associated DMPs comparing sPE_Type2 versus sPE_Type1 samples, with sPE_Type1 as the reference. Top 10 significant CpGs are annotated with corresponding gene symbols. (d) Pathway enrichment analysis of subtype-associated DMPs from KEGG database. (e) Comparison of HOME-estimated placental cell-type proportions in term, GA-matched preterm and the two sPE subtypes. Wilcoxon rank-sum tests were used for subtype comparison. All pair-wise comparisons are significant (P<0.05), except those labeled as ns. ns: not significant. (f) Evaluation of an elastic-net based classification biomarker model constructed from in-house type 1 vs type 2 sPE samples (training n = 136; testing n = 33; 8:2 split with five-fold cross-validation) by receiver operating characteristic (ROC) curve, using in-house training and in-house testing datasets as labeled. (g) External validation of the sPE epigenetic subtype classifier built in (f) by six independent placental methylation cohorts. The classifier predicts two placental methylation subtypes in external datasets. PCA projection shows subtype assignments across combined datasets. (h) External validation of SGA risk in inferred subtype 2 samples in GSE98224, measured by ROC analysis.

We next examined CpG methylation changes in Subtype 2 relative to Subtype 1 as baseline (**Figure 4c**), identifying 20 hypermethylated and 59 hypomethylated CpGs (1 and 7, respectively, in promoter-proximal TSS1500 regions). Hypermethylated CpGs included a gene-body site in JARID2, a Polycomb (PRC2) cofactor enriched in progenitor cells^38^, and a TSS1500 site in TFAP2B, a CTB-identity transcription factor^39^ (**Figure 4c; Supplementary Table 5**). Hypomethylated CpGs at 5′UTR and TSS1500 regions included angiogenesis-associated genes (SMAD9, RCAN2) and the imprinted growth suppressor GRB10^40^. Pathway enrichment of Subtype 2 over Subtype 1 (**Figure 4d**) implicates stem-cell pluripotency signaling (SMAD9, JARID2) and the repressed mTOR signaling axis (GRB10, LPIN1), suggesting increased stemness, repressed metabolic adaptation, and trophoblast stress in subtype 2.

Compared to differential methylated CpGs, cell composition variation appears to be more influential between the two methylation subtypes (**Figure 4e)**. sPE subtype 2 shows expansion of CTBs and EVTs compartments, but reduction of SCTs, endothelial, fibroblast and Hofbauer cells, compared with subtype 1 and pre-term controls (P < 0.05). This CTB/EVT-increasing and SCT-decreasing profile in subtype 2 shows that trophoblast maturation is hampered, leading to progenitor-like and invasive trophoblast accumulation and CTB to SCT differentiation inhibition^41^. Reduction of other cell types in the villous core further supports impaired villous remodeling and reduced fetal-maternal exchange capacity^13^, providing the cellular basis for the increased SGA risk in subtype 2 of sPE. By contrast, sPE subtype 1 presents the opposite pattern with lower CTB but higher SCT proportions than pre-term controls, demonstrating a hypermature villous profile. Thus, methylation stratifies clinically similar sPE into a hypermature placenta villous type (Subtype 1) and an immature placenta villous type (Subtype 2), which may explain the difference in SGA risks.

We next validated the robustness of these two sPE subtypes using external cohorts. We first built a elastic-net classifier on sPE subtype-discriminative CpGs from HiBR placenta cohort, which achieves high accuracy (AUC = 0.997 in training data and 0.996 in hold-out testing of the same cohort (**Figure 4f**). Classifier features include developmental transcription factors and placental transport genes including LHX4, GCM2, TFAP2B and SLC2A3^42–44,45^, showing the developmental basis of sPE subtypes (**Supplementary Table 6**). We then applied the classifier to identify the two-subtypes across six independent placental methylation datasets^10,46–50^ comprising 150 preeclampsia cases (**Figure 4g**). In the external cohort GSE98224 which has fetal growth information, the same classifier further predicts SGA, which is preferably associated with inferred sPE Subtype 2, with an accuracy of AUC = 0.701 (**Figure 4h**). Therefore, the sPE subtypes represent reproducible and different placental molecular states. They not only preserve the epigenetic biomarker signals across datasets but are also associated with different fetal growth risks clinically.

### Placenta clock modeling shows different rates of gestational age acceleration in the two sPE subtypes

Accelerated placental aging has been speculated as a feature of preeclampsia, with epigenetic clocks showing that PE placentas deviate from the normal maturation trajectory and appear older than their chronological gestational age (GA) ^51,52^. To accurately quantify aging acceleration within the sPE subtypes, we harmonized a collection of 692 uncomplicated placentas from additional 9 studies, whose GAs range from 8 to 41 weeks (**Figure 5a; Supplementary Table 7**). We trained a placental gestational-age (GA) clock by elastic-net regression on 80% of the samples, which predicts the GA accurately in the 20% held-out samples (PCC = 0.98; **Figure 5b**). The model includes 235 GA associated CpG features (**Supplementary Table 8**), with the top 20 features shown in **Figure 5c**. Leading CpG loci map to genes governing trophoblast lineage development (SATB1^53^), decidualization and implantation (HOXA10^54^, TINAGL1^55^), and programmed cell death (DFFA^56^, PEBP1^57^), reflecting the placental developmental processes.

**Figure 5.**
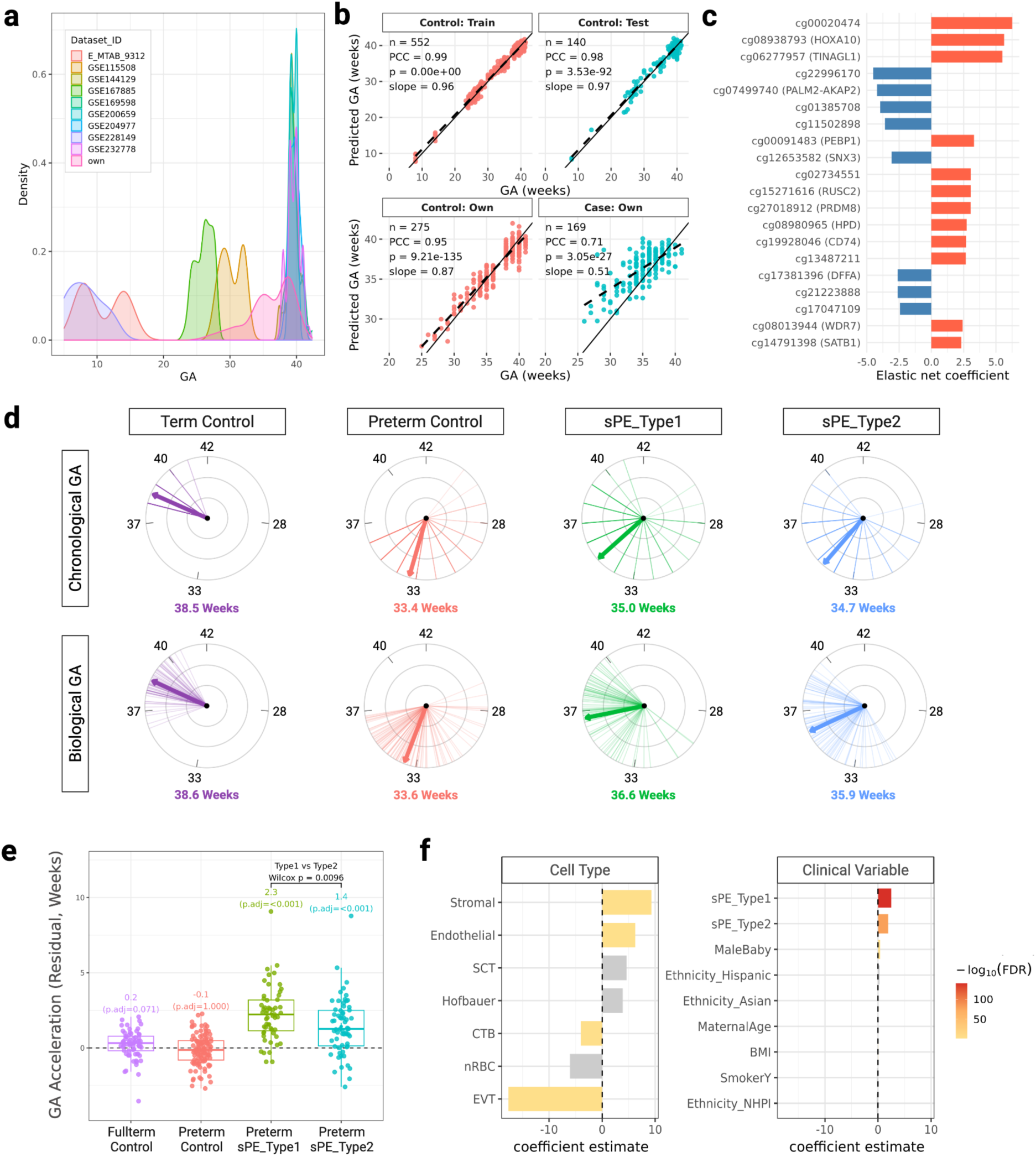
Placental epigenetic clock modeling shows that the two sPE epigenetic subtypes have accelerated placenta aging, but at different rates. (a) GA distributions across nine public and in-house placental methylation cohorts (n=692) were used for placental epigenetic clock construction. (b) Performances of the Elastic-net normotensive placental gestational age clock in training, hold-out testing, as well as our own control and sPE cohort. PCC is the metric used to evaluate the correlation between predicted biological GA (y-axis) and chronological GA (x-axis) of placentas. (c) Top 20 elastic-net CpG features contributing to the placental epigenetic clock model. (d) Circular placenta GA clock visualization comparing chronological GA (top row) and DNAm-predicted biological GA (bottom row) across the four groups: term control, preterm control, sPE subtype 1 (sPE_Type1), and sPE subtype 2 (sPE_Type2). Each radial line represents one sample, positioned according to GA on a 20-42 week circular scale. The colored thick arrow denotes the group means GA for each condition, with the mean GA value shown below each clock. (e) Box plots of placenta GA acceleration values in four groups. From left to right: term control, preterm controls, preterm sPE_Type1, and preterm sPE_Type2. Placental GA acceleration is defined as the residual from the regression of DNAm-predicted biological GA on observed chronological GA. Adjusted P values indicate one-sample tests comparing GA acceleration with zero after Bonferroni correction. The P value above the bracket indicates the Wilcoxon rank-sum test comparing preterm sPE_Type1 and preterm sPE_Type2. (f) Associations between placental cell-type proportions, clinical covariates, and GA acceleration estimated by multivariable regression. Significant coefficients are colored, whereas non-significant estimates are shown in grey.

We applied the constructed placenta GA clock model in our control term and preterm placentas, neither of which shows strong evidence of GA acceleration relative to their chronological GAs (term, +0.2 weeks, P.adj = 0.071; preterm, -0.1 weeks, P.adj = 1.000; **Figure 5d,e**). In contrast, both sPE subtypes show GA acceleration, with predicted biological GA greater than their chronological GA. sPE Subtype 1 shows significantly greater GA acceleration than sPE subtype 2 (+2.3 versus +1.4 weeks; Wilcox P=0.0096), despite having lower SGA burden than subtype 2 (**Figure 5d,e; Supplementary Figure 6**). Thus, GA acceleration does not necessarily scale with fetal growth restriction.

We next explored the major factors contributing to the difference in GA acceleration in the two subtypes, regressing GA acceleration (y-value) by cell-type proportions, subtypes and other clinical covariates (**Figure 5f**). GA acceleration is significantly positively associated with stromal and endothelial cells in the villous core, but negatively with CTBs and EVTs (FDR<0.05). Their coefficients are much larger than the subtype 1 and subtype 2 identities (stromal = +9.28, endothelial = +6.23, CTB = −4.02, EVT = −17.66, versus subtype 1 = +2.52, subtype 2 = +1.92). As subtype 1 has significantly lower CTB but much more stroma and endothelial cells (**Figure 4e**), this explains why it has more accelerated GA than subtype 2. This also aligns with prior studies indicating that placentas with higher stromal content and lower proportions of progenitor CTBs and invasive EVTs are biologically older^58,59^. Together, these findings show that accelerated placental aging reflects villous and trophoblast maturation states.

### Multi-omics integration validates the two sPE subtypes and their molecular mechanisms

Proteomics, unlike epigenomics, provides direct measurements on sPE function readouts at protein level. We thus conducted DIA proteomics assays on a subset of 158 placentas (78 sPE, 80 controls) which are propensity-matched. The proteomic data alone recapitulate established sPE pathology, including elevated FLT1 and PAPPA2, activation of complement, coagulation, and VEGFA-VEGFR2 signaling, and reduced oxidative phosphorylation (**Figure 6a,b**).

**Figure 6.**
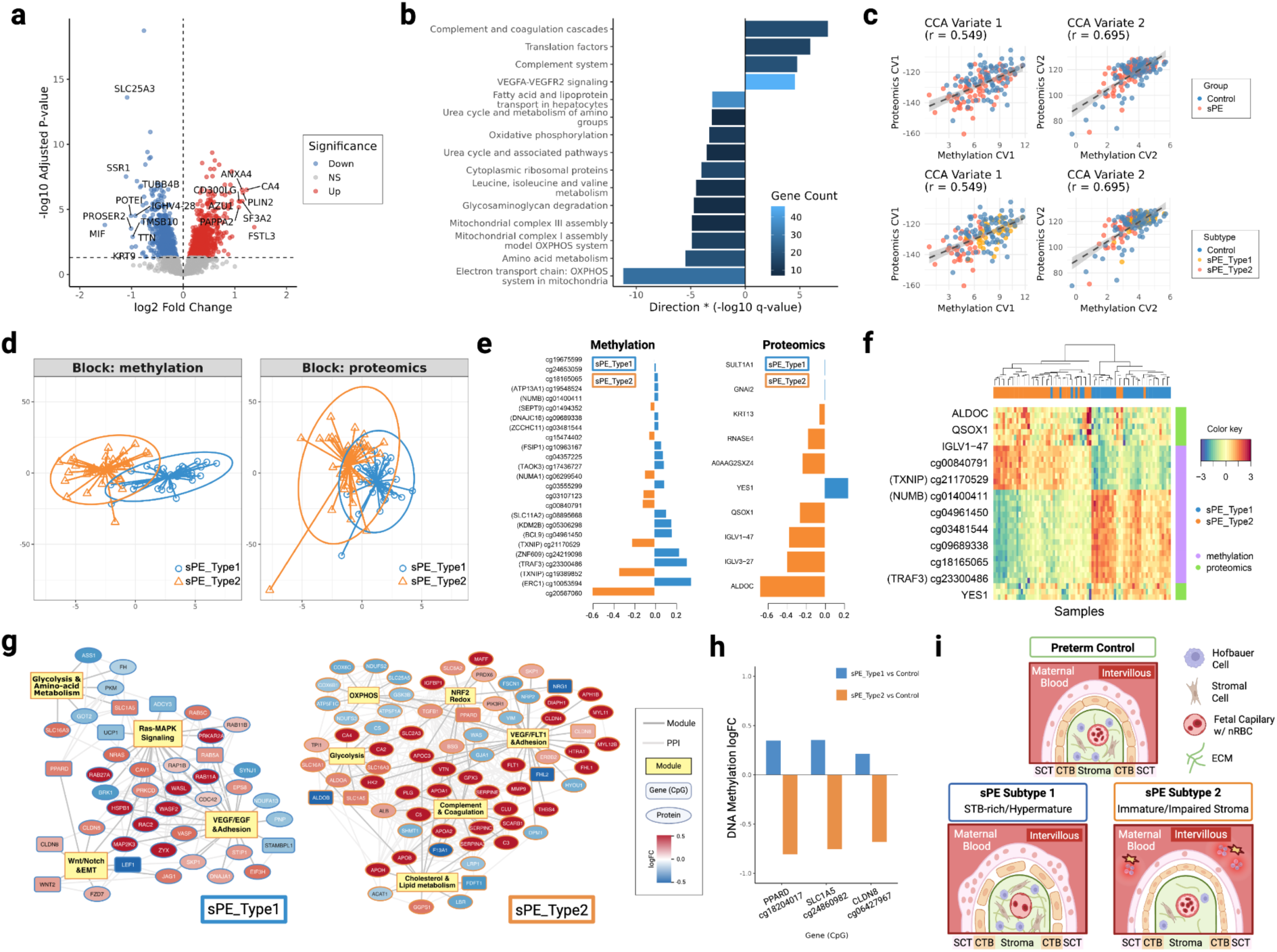
Proteomic assays validate the two sPE epigenetic subtypes. (a) Volcano plot of differential proteins in sPE (n=78) versus propensity-matched controls (n=80). (b) Proteomic pathway enrichment analysis results, pathways shown are after FDR=0.05 thresholding. (c) Integration of methylation and proteomic profiles by sparse canonical correlation analysis (sCCA) identifies two major cross-modal axes of variation (CV). Pearson correlation between paired latent variables (r) is reported. (d) Projection of sPE subtypes in methylation and proteomics latent spaces demonstrates coordinated subtype separation across omic layers. (e) Top methylation and proteomic features contributing to the first latent component separating sPE subtypes. (f) Integrated heatmap of top cross-modal subtype-associated methylation and proteomic features. CpGs with known gene symbols are annotated. (g) Subtype-specific CpG-protein interaction networks derived from cross-modal correlated features and annotated by PPI enrichment. Nodes represent CpG or proteins, edges indicate Protein-Protein Interaction (PPI) connections, and node color denotes log fold change relative to controls. (h) Common CpGs present in the PPI networks of both subtypes in (g). logFC comparison for each compared with all propensity-matching controls is reported. (i) Conceptual model showing the distinct molecular mechanisms of each sPE subtype compared to the matching preterm control.

We integrated the methylation and proteomics by sparse CCA, which identifies two paired methylation-protein axes: one capturing cytoskeletal remodeling and trophoblast invasion (CV1), the other nuclear, mitochondrial and epigenetic regulation (CV2) (canonical correlations 0.549 and 0.695; **Figure 6c; Supplementary Table 9**). Supervised multi-omics integration using DIABLO shows proteomic signatures classify the methylation-defined subtypes with 88.2% accuracy, indicating that the epigenetic subtypes are well-accompanied by distinct functional proteomic states (**Figure 6d-f**). Subtype 1 is marked by the proliferative Src-family kinase YES1, whereas subtype 2 has a high level of hypoxia-inducible glycolytic enzyme ALDOC^60^ together with oxidative-stress-associated oxidase QSOX1^61^. Additionally, subtype 2 has significantly elevated immunoglobulin light-chain proteins (IGLV1-47, IGLV3-27), linking subtype 2 to immune-driven form of placental pathology (**Figure 6e,f**).

We next visualized the integrated highly correlated CpG and proteins, in the context of PPI networks (**Figure 6g; Supplementary Table 10**). The two sPE subtypes present drastically different pathogenic programs. Subtype 1 forms a fusion-competent villous-remodeling network, in which glycolytic and amino-acid metabolism is linked to Ras-MAPK signaling, VEGF/EGF-dependent adhesion and Wnt/Notch/EMT remodeling. Cytoskeletal regulators such as CDC42, RAC2 and WASF2 are connected in these modules, consistent with active trophoblast differentiatiosn and SCT-rich villous state (**Figure 6g; Supplementary Table 10**). By contrast, subtype 2 forms a broader injury network spanning anti-angiogenic VEGF/FLT1 signaling, complement and coagulation, hypoxia-linked metabolic remodeling marked by glycolysis and reduced OXPHOS, NRF2 redox stress and cholesterol and apolipoprotein metabolism linked to both endothelial dysfunction and immune activation **(Figure 6g; Supplementary Table 10)**. Three CpGs in the promoter regions of PPARD, SLC1A5 and CLDN8 show up in PPI networks of subtype 1 and 2 both. Interestingly they have opposite methylation shifts between the two subtypes **(Figure 6h**), linking subtype-specific methylation to lipid-metabolic differentiation, amino-acid transport/syncytin-receptor biology and epithelial junction remodeling^45,62^. Together, multi-omics integration confirmed two divergent sPE placental programs: a hypermature Subtype 1 defined by accelerated villous maturation and syncytial differentiation and a growth-restricted Subtype 2 defined by impaired trophoblast maturation, hypoxia and increased immune-activation stress. **(Figure 6i)**.

## Discussion

Severe preeclampsia (sPE) is currently managed as a single disorder, yet this clinical label may conceal biologically distinct placental disease programs that converge on maternal hypertension and end-organ dysfunction. Defining these hidden placental states is therefore essential for moving beyond one-size-fits-all obstetric management toward precision obstetric medicine, enabling improved risk stratification, prenatal monitoring, and mechanism-guided intervention. Our findings provide strong evidence that sPE comprises multiple molecularly and cellularly distinct placental subtypes with divergent developmental trajectories, biological mechanisms, and clinical outcomes. These results support a reclassification of sPE based on underlying placental biology rather than clinical presentation alone and establish a framework for biologically informed diagnosis and targeted therapeutic development.

Here, we show that clinically similar sPE pregnancies can arise from distinct placental disease programs associated with divergent fetal growth outcomes. In a clinically matched, multi-ethnic cohort of 444 placentas from HiBR, integrated DNA methylation and proteomic profiling identified two reproducible sPE subtypes that were largely indistinguishable by conventional clinical characteristics yet differed markedly in placental composition, developmental state, and fetal growth risk. Subtype 1 was characterized by a CTB-reduced/SCT-rich, hypermature villous phenotype accompanied by greater placental epigenetic age acceleration, but slightly higher risk of small-for-gestational-age birth than matching controls. In contrast, subtype 2 exhibited expansion of CTB and EVT populations, reduction of SCTs and cells in the villous core including Hofbauer cells and fibroblasts, a markedly higher incidence of small-for-gestational-age birth but less placental epigenetic age acceleration, compared to subtype 1. Together, these findings suggest that a common maternal syndrome can emerge from distinct placental pathophysiological states, providing a biological framework for stratifying sPE beyond its clinical presentation.

Resolving molecular heterogeneity in sPE first requires overcoming a longstanding challenge in placental epigenomics: pervasive cellular heterogeneity, further compounded by differences in gestational age. Bulk placental methylation profiles capture both cell-intrinsic regulatory variation and shifts in tissue composition, making it difficult to distinguish disease-associated mechanisms from developmental remodeling. This challenge is particularly acute in trophoblast lineages, where CTBs, SCTs, and EVTs exist along a continuous developmental trajectory yet perform distinct biological functions. Existing deconvolution approaches, including single-cell-informed frameworks such as EpiSCORE^30^, placenta-specific reference panels such as PlaNET ^63^ and Campbell et al. ^64^ have advanced the field but do not fully resolve trophoblast subtype architecture, particularly EVTs, which are central to spiral artery remodeling and placental vascular adaptation.

To address this limitation, we developed HOMED, a hierarchy-aware deconvolution framework that leverages complementary information from placental single-cell transcriptomes, purified-cell methylomes, and paired bulk methylation profiles to resolve trophoblast lineage complexity. This approach enables separation of disease-associated methylation changes from cellular compositional remodeling at trophoblast-subtype resolution. Applied across the HiBR cohort, HOMED reduced genomic inflation by approximately 170-fold, demonstrating that a large fraction of methylation signals previously attributed to preeclampsia likely reflect variation in cell composition and gestational age rather than disease-specific epigenetic regulation. Importantly, this increased resolution did more than improve statistical calibration. It revealed that the newly identified sPE subtypes are distinguished by fundamentally different trophoblast architectures, including divergent balances of CTBs, SCTs, EVTs and villous-core cell populations. These biological differences were largely obscured using coarser deconvolution strategies, highlighting how resolving cellular heterogeneity is essential for uncovering disease substructure. More broadly, our findings suggest that accurate interpretation of placental methylation studies requires modeling trophoblast lineage complexity, and establish HOMED as a framework for disentangling cellular and regulatory mechanisms in heterogeneous tissues.

Multi-omic integration places the methylation-defined subtypes into a broader framework of placental disease heterogeneity. Previous molecular studies of preeclampsia have implicated trophoblast dysfunction, anti-angiogenic signaling, inflammation, and metabolic stress, but these processes are typically interpreted as components of a single placental syndrome ^9,66^. Our findings suggest instead that these canonical pathways are differentially configured across biologically distinct forms of sPE. Subtype 1 is characterized by a CTB-reduced/SCT-rich hypermature villous state associated with coordinated remodeling of trophoblast fusion, epithelial adhesion, cytoskeletal organization, and metabolic adaptation. In contrast, Subtype 2 exhibits features of impaired placental maturation, including expansion of less differentiated CTB populations, depletion of SCT, and loss of villous-core cell populations. These cellular alterations are accompanied by activation of FLT1-associated vascular stress pathways, complement and coagulation signaling, and hypoxia-related metabolic remodeling. Together, these results indicate that clinically similar sPE pregnancies can arise through distinct placental pathophysiological trajectories: a hypermature adaptive program in Subtype 1 and a maturation-arrested, thrombo-inflammatory, growth-restricted program in Subtype 2.

Our findings also challenge the prevailing view that placental gestational-age acceleration primarily reflects cumulative stress, premature senescence, or trophoblast dysfunction ^51,52,67^. Despite exhibiting greater disruption of trophoblast differentiation and a substantially higher risk of fetal growth restriction, Subtype 2 (+1.4 weeks) showed weaker placental age acceleration than Subtype 1 (+2.3 weeks). This apparent paradox was largely explained by cellular composition: age acceleration was positively associated with mature core compartments including stroma and endothelial cells and negatively associated with the accumulation of progenitor CTBs and maladaptively expanded EVTs. These observations suggest that placental epigenetic aging is not a unidirectional measure of disease severity, but rather a state-dependent readout of placental developmental organization. Within sPE, faster GA acceleration appears to mark a hypermature, stress-adapted villous program, whereas slower GA acceleration is associated with impaired trophoblast maturation and persistence of less differentiated cellular states. More broadly, our results indicate that placental epigenetic clocks capture distinct developmental trajectories rather than a simple continuum of pathological burden (as shown by SGA). Interpretation of placental aging therefore requires consideration of cellular architecture and disease subtype, particularly in heterogeneous disorders such as sPE.

In summary, our findings redefine sPE as a heterogeneous placental disorder composed of distinct molecular and cellular disease states. By resolving this previously hidden heterogeneity, we establish a foundation for subtype-specific biomarkers, prenatal detection, and precision approaches to the management of preeclampsia.

## Methods

### HiBR placenta cohort

Placental tissue samples were obtained from the RMATRIX Hawaii Biorepository (HiBR) under institutional review board approval. As reported in other studies^68^, the Hawaii Biorepository (HiBR) contains over 9000 triads of maternal blood, fetal blood, and placental tissue from births at Kapi‘olani Medical Center for Women and Children (KMCWC), linked to clinical data obtained from the electronic medical records. The HiBR was supported by National Institute on Minority Health and Disparities and was approved by the Western Institutional Review Board (WIRB Study Number #1107593). The repository obtains informed consent from postpartum participants for the donation of placental tissue, umbilical cord tissue, and residual umbilical cord and maternal blood specimens. After clearance by the medical team, placentas were transported to the HiBR in phosphate buffer saline (PBS). The HiBR sample collection protocol consisted of incising 50 g of placental tissue from 2 cotyledons obtained mid-region of the villous parenchyma, midway between the umbilical cord insertion and the placental margin. Selected cotyledons were excised, cut into smaller pieces, placed into a 50 mL polyethylene (PET), and placed into a −80 °C freezer for long-term storage without any additive or cryopreservation solutions.

The study includes a total of 444 human placenta tissues at the chorionic villous location, with 169 placentas from pregnancies complicated by severe preeclampsia (sPE) meeting established diagnostic criteria, 169 normal term controls, and 106 preterm controls (**Figure 1**). Severe preeclampsia is defined as ICD-9 criteria for high blood pressure and significant proteinuria with potential organ involvement (e.g., liver/kidney) or symptoms like vision changes and abdominal pain (ICD-9 code 642.5).

### Propensity score-based sample matching

To further mitigate confounding, propensity score (PS) matching was performed in R using the MatchIt (v4.7.2)^69,70^ package for matching implementation. Propensity scores were estimated using logistic regression with relevant maternal and pregnancy covariates, incorporating maternal age, BMI, parity, ethnicity, smoking status, and gestational age. One-to-one nearest-neighbor matching without replacement was performed under the average treatment effect on the treated (ATT) framework based on the logit-transformed propensity score. Matched sPE-control pairs (169 controls and 169 sPE cases) were subsequently used for downstream analyses.

### DNA isolation and genome-wide DNA methylation profiling

Fresh-frozen placental tissues were minced with disposable scalpels and homogenized on ice using disposable micro-pestles. The protein components of the tissue preparation were removed by incubating with Proteinase K at 56°C for one hour. Genomic DNA was extracted from all the tissue samples using the AllPrep DNA/RNA Kit (Qiagen), following the manufacturer’s protocol. DNA quantification and quality check were performed prior to downstream processing. Samples with RNA or protein impurities were further purified to get high-quality DNA samples for processing.

Genome-wide DNA methylation profiling was performed using the Illumina Infinium MethylationEPIC BeadChip, which interrogates over 850,000 CpG sites across the human genome. DNA quality was evaluated using NanoDrop spectrophotometry and PicoGreen fluorescence assays to ensure suitability for array-based profiling. Bisulfite conversion was carried out using the EZ DNA Methylation Kit (Zymo Research), and 100-250 ng of bisulfite-converted DNA per sample was hybridized to the EPIC BeadChip according to the manufacturer’s instructions. Arrays were scanned using Illumina iScan instrumentation, and raw intensity data were generated in IDAT format for downstream analysis.

### Methylation data preprocessing

All analyses were performed in R (v4.3.2). The methylation preprocessing pipeline is summarized in **Supplementary Figure 1**, following established procedures^71^. Sample sex was verified using minfi (v1.48.0) getSex() function^72^. Raw IDAT files were imported via ChAMP (v2.16.2) with background correction^73^. Probes failing detection *P* > 0.05, sex-chromosome probes, SNP-containing probes, and low-quality probes were excluded. BMIQ normalization corrected probe-type bias. Batch effects were adjusted using ComBat, confirmed by SVD analysis^74^. Cross-hybridizing probes were removed per ExperimentHub annotations (EH3129). Beta values were converted to M-values using lumi (v3.1.4)^75^. The final dataset contains 696,557 CpGs and 444 samples.

### Overview of HOMED (Hierarchically Optimized MEthylation Deconvolution)

For placental cell-type deconvolution from the bulk DNA methylation data, we created HOMED (Hierarchically Optimized MEthylation Deconvolution), a hierarchical framework for estimating cell-type proportions. The HOMED placenta reference panel was constructed by integrating three data sources using the HOMED_Reference() function: (1) FACS/MACS-sorted placental methylation profiles, (2) the placental single-cell atlas, (3) paired bulk RNA and DNA methylation datasets (**Figure 3a**).

1. *FACS/MACS-sorted placental purified cell data*. Purified placental methylation profiles for seven major cell types were assembled from public sources as listed in **Supplementary Table 2**: nRBC (FlowSorted.CordBloodCombined.450k52)^76^, EVT (GSE6088553)^77^, CTB and SCT (GSE27169754, GSE15952655)^64,63^, and stromal, Hofbauer, endothelial (GSE15952655)^63^. Datasets were normalized using noob in minfi (v1.48.0)^78^, batch-corrected using ComBat in sva (v3.50.0)^74^, restricted to shared CpGs and concatenated (**Supplementary Figure 2a-b**). Gestational DMR probes^71^ were removed to limit GA confounding (**Supplementary Figure 2c**).
2. *Placental single-cell atlas.* A high-resolution placental single-cell transcriptomic atlas was assembled by integrating six publicly available control placental single-cell RNA-seq datasets^13,79–83^. Data were processed using standard normalization, harmony integration, dimensionality reduction, and clustering workflows in Seurat^84^. Cell identities were assigned based on canonical marker genes and curated annotations, yielding 13 cell types. This atlas served as a reference for bulk RNA deconvolution.
3. *Paired bulk RNA and DNA methylation datasets.* Two placenta datasets with matched RNA and methylation from the same AllPrep specimens were used (GSE98224, GSE7337724; n = 111)^10,49^. RNA expression was deconvolved against the placenta single-cell atlas with DWLS^85^ and SCDC^86^. SCDC result was selected as pseudo-ground-truth proportions as it yielded the most biologically consistent estimates (higher trophoblast fraction, biologically plausible Hofbauer/nRBC/EVT ranges; **Supplementary Figure 3**).

### Cell type deconvolution using HOMED

HOMED optimizes hierarchy-aware DNAm reference libraries using a two-stage IDOL-based framework^87^ to resolve developmentally related trophoblast subtypes while preserving major placental cell-type separation (**Figure 2a**). The HOMED framework consists of six major steps: (1) integrate sorted-cell methylation profiles into an initial reference; (2) identify candidate cell-type-specific CpG markers using one-versus-rest differential methylation analysis; (3) estimate pseudo-ground-truth cell proportions from paired bulk RNA-seq data using SCDC; (4) iteratively optimize CpG selection probabilities by evaluating deconvolution performance against the RNA-derived proportions, retaining CpG sets that improve RMSE and PCC; (5) generate optimized hierarchical reference libraries for each layer; and (6) apply the final reference to deconvolve new bulk DNAm samples.

During optimization (steps 3-4), each candidate signature matrix was evaluated by deconvolving training methylation samples using EpiDISH constrained projection (CP) (v2.18.0) ^30^, and CpGs contributing to improved deconvolution accuracy were assigned higher sampling probabilities in subsequent iterations. Optimization was performed for a predefined number of iterations while retaining the best-performing CpG library according to RMSE improvement thresholds. Optimized CpG signature matrices from both hierarchical layers were subsequently normalized into a unified reference and applied to 444 bulk samples using the HOMED_Estimate() function.

### Validation of HOMED method

To evaluate HOMED’s deconvolution accuracy under a controlled ground truth, we constructed pseudo-bulk methylation samples from purified cell-type reference profiles with ground-truth cell proportion modeled from Dirichlet distributions, and compared the estimated cell-type proportions to the known mixing weights.

#### Pure cell sampling

Let K denote the number of cell types included in the reference panel (Hofbauer cell, nRBC, endothelial, stromal, EVT, SCT, and CTB; K=7). For each cell type k∈{1,…,K}, we randomly drew n pure methylation profiles (default n=3) from the reference pool, yielding a pure cell matrix:

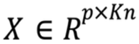, where p is the number of CpG sites and each column x_k,j_ (j=1,…,n) corresponds to the j-th pure sample of cell type k.

#### Mixture weight generation

For each simulated bulk sample m∈{1,…,M}, the within-cell-type contributions across the n replicate pure cell profiles of cell type k were drawn from a Dirichlet distribution,

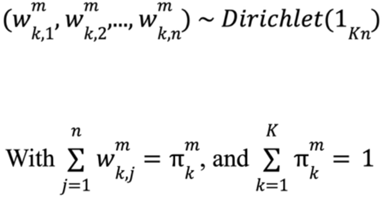

Where 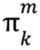 is the summative true proportion of cell type k in each sample m. Stacking across cell types and samples produced the weight matrix: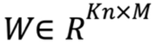.

#### Pseudo-bulk construction

The simulated bulk methylation beta matrix was generated by a linear mixture of the pure cell profiles: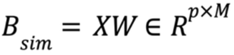.

#### Performance metrics

HOMED was applied to B_sim_ to obtain estimated proportions 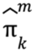 and compared with the ground-truth proportion 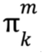. Pearson correlation coefficient (PCC) and Root Mean Square Error (RMSE) were used to quantify the agreement between the estimated and ground-truth proportions.

The details of HOMED benchmarking is reported elsewhere^32^. Among PBMC, lung and placenta datasets, HOMED consistently improved deconvolution accuracy compared with existing single-cell RNA-Seq guided DNAm deconvolution methods, EpiSCORE and scDeconv. HOMED also incorporates a batch-based optimization strategy that substantially reduces computational runtime while maintaining high deconvolution performance^32^.

### Source of variance analysis and confounding adjustment

SOV analysis^11,71^ across clinical variables and deconvolution-derived cell types identified adjustment covariates. As cell fractions are compositional (sum-to-one), nRBC was dropped for identifiability^11^. Per-variable ANOVA across CpGs retained variables with F > 1; smoking was included as a standard EWAS covariate^11,88^. Final covariates included endothelial, Hofbauer, stromal, EVT, SCT, and CTB proportions, gestational age, fetal sex, gestational diabetes, ethnicity, pre-pregnancy BMI, and smoking status.

### Differential methylation analysis and genomic inflation correction

Differential analysis was performed using the limma (v3.58.1)^89^, adjusting for clinical confounders and HOMED estimated cell proportion listed above. Test-statistic inflation was corrected with bacon^35^ (post-adjustment λ = 1.07; **Supplementary Figure 4a-b**). Benjamini-Hochberg FDR was applied to bacon-corrected statistics. Probes were annotated with IlluminaHumanMethylationEPICanno.ilm10b4.hg1967^90^. CpG region information was examined (**Supplementary Figure 4c**). Pathway enrichment used enrichR (v3.2)^91^ by aggregating the CpG sites to the gene level (Reactome, WikiPathways; BH-adjusted q < 0.05).

### sPE methylation subtyping

sPE subtypes were defined by unsupervised clustering of placental DNA methylation profiles. Beta values were first adjusted for clinical confounders (gestational age, gender, BMI, gestational diabetes, ethnicity, maternal age and smoking status) to minimize non-biological variation before subtype discovery. CpG sites were ranked by median absolute deviation (MAD) across samples, and the 50,000 most variable sites were retained for clustering. Consensus clustering (K-means, NMF, spectral^92^) converged on k = 2 (silhouette-optimal, concordant across all three methods; **Supplementary Figure 5**). Subtypes were labeled by SGA frequency (Subtype 2 = higher SGA). Clinical and demographic statistics for identified sPE subtypes are reported in **Supplementary Table 4**. Differential methylation analysis between subtypes was performed using limma (v3.58.1)^89^, adjusting for clinical confounders and HOMED estimated cell proportion, comparing Subtype 2 to Subtype 1 with Subtype 1 as the reference group. Pathway enrichment was done using enrichR (v3.2)^91^ KEGG database.

### External validation of sPE subtypes

For external validation, the in-house discovery data were harmonized with six public cohorts (GSE98224^10^, GSE100197^46^, GSE125605^47^, GSE57767^48^, GSE73375^49^, GSE75196^50^; 150 preeclampsia cases total), followed by quantile normalization in preprocessCore (v1.64.0)^93^ and ComBat^74^ batch correction to reduce cross-study technical variation.

Subtype-discriminative CpGs were used to train an elastic-net classifier (caret (v6.0-94)/glmnet (v4.1-8))^94,95^ on in-house sPE samples only with an 80/20 train/test split. Hyperparameters (α ∈ [0,1]; λ over 30 log-spaced values) were tuned by 5-fold cross-validation repeated three times, optimizing CV AUC. Non-zero features for the sPE subtype classifier were reported in **Supplementary Table 6**. The finalized sPE classifier was then applied to the harmonized six public datasets to evaluate the robustness of methylation-defined sPE subtypes. SGA predictive performance was assessed by the area under the receiver operating characteristic curve (AUROC) in GSE98224^10^, the validation cohort with available small-for-gestational-age (SGA) status.

### Placental epigenetic aging clock model

A GA clock was trained on 692 normotensive controls from ten datasets (nine public datasets and our in-house data). Nine datasets (E_MTAB_9312^96^, GSE115508^97^, GSE144129^98^, GSE167885^99^, GSE169598^100^, GSE200659^101^, GSE204977^102^, GSE228149^103^, GSE232778^104^) were curated by the DREAM Placental Clock DNA Methylation Challenge^105^ (https://synapse.org/Synapse:syn59520082) (**Supplementary Table 7**). Samples were split into training and test sets by stratified sampling across GA quantiles, with an 80/20 train/test split. Gestational-age-associated candidate CpGs were first filtered by Spearman correlation between beta values and gestational age, and CpGs with |ρ| > 0.5 were further tested using limma on M-values while adjusting for fetal sex and dataset. CpGs at FDR < 0.05 were retained as the input feature set for elastic net regression. An elastic net model was fitted on the training set using cv.glmnet (Gaussian family, α = 0.5) with 10-fold cross-validation and mean squared error as the selection criterion from the *glmnet* package^95^. The optimal regularization parameter was defined by lambda.min, and 235 CpGs with non-zero coefficients at this penalty were retained in the final model (**Supplementary Table 8)**.

The fitted clock was applied to control training samples, held-out control test samples and in-house sPE placentas. Predictive performance was assessed by Pearson correlation between chronological GA and methylation-predicted GA. GA acceleration was calculated as the residual from a control-calibrated linear model relating predicted biological GA to chronological GA, as previously described by Lee et al.^51^ Group-level deviation from zero GA acceleration was tested using one-sample Wilcoxon signed-rank tests, with Bonferroni adjustment.

Associations among placental GA acceleration, inferred cell-type composition, and clinical variables were evaluated using multivariable linear regression. To avoid collinearity from compositional cell-fraction data, each model included one cell type at a time:

𝐴𝑐𝑐𝑒𝑙𝑒𝑟𝑎𝑡𝑖𝑜𝑛 ∼ 𝐶𝑒𝑙𝑙𝑇𝑦𝑝𝑒_!_ + 𝐺𝑟𝑜𝑢𝑝 + 𝑆𝑒𝑥 + 𝐵𝑀𝐼 + 𝐸𝑡ℎ𝑛𝑖𝑐𝑖𝑡𝑦 + 𝑀𝑎𝑡𝑒𝑟𝑛𝑎𝑙𝐴𝑔𝑒 + 𝑆𝑚𝑜𝑘𝑖𝑛𝑔

Since each model included one cell type at a time, clinical covariate effects were summarized across models using fixed-effect inverse-variance weighting, with significance assessed by two-sided Wald tests.

### Placenta proteomics experiment

A total of 158 propensity-score matched placenta samples (control N=80, sPE N=78) were selected from the methylation cohort. Approximately 100 mg of freshly frozen placenta tissue from each sample was used for analysis. Clarified protein lysates were solubilized with 1X RIPA, normalized across 158 placenta samples using BCA quantification, reduced with TCEP, and alkylated with Iodoacetamide. Protein (70ug) was digested with sequencing grade, modified porcine trypsin (Promega) using S-Trap 96-well plates following the protocol from the manufacturer (ProtiFi).

Tryptic peptides were then separated by a reverse phase Ion-Opticks-TS analytical column (25 cm x 75 um with 1.7 um C18 resin) supported by an EASY-Spray nano-source and stabilized with a Heater THOR Controller (Ion-Opticks) at 60°C. Peptides were trapped and eluted from a (PepMap Neo, 300um x 5mm Trap) using a Vanquish Neo UHPLC nano system (Thermo Scientific), which kept the samples at 11°C before injection. Peptides were eluted at a flow rate of 0.350uL/min using a 35 min gradient from 98% Buffer A:2% Buffer B to 94.5:5.5 at 0.1 minutes to 56:44 at 27.1 minutes followed by a column wash of 45:55 at 29.7 minutes to 1:99 at 35 minutes followed by equilibration back to 98:2. Eluted peptides were ionized by electrospray (2.5 kV) followed by mass spectrometric analysis on an Orbitrap Astral mass spectrometer (Thermo). Precursor spectra were acquired from 380-980 Th, 240,000 resolution, normalized AGC target 200%, maximum injection time 3 ms. DIA acquisition on the Orbitrap Astral was configured to acquire 199, 3 Th window from 380-980 Th, 25% HCD Collision Energy, normalized AGC target 100%, maximum injection time 3 ms. Fragment (MS2) scan range from 150-2000 Th with an RF Lens (%) set to 40.

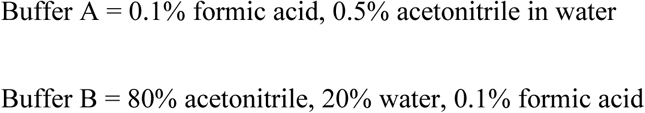

### Proteomics data preprocessing and analysis

Following the proteomics data acquisition, data were searched using Spectronaut (Biognosys version 20.3) against the UniProt *Homo sapiens* database (Proteome ID: UP000005640, Taxon ID: 9606, 4^th^ version of 2025) using the directDIA method with an identification precursor and protein q-value cutoff of 1%, generate decoys set to true, the protein inference workflow set to Quant 2.0, inference algorithm set to IDPicker, quantity level set to MS2, cross-run normalization set to false, and the protein grouping quantification set to median peptide and precursor quantity. Fixed Modifications were set to Carbamidomethyl (C) and variable modifications were set to Acetyl (Protein N-term), Oxidation (M).

Protein MS2 intensity values were assessed for quality using ProteiNorm^106^. The data were normalized using Cyclic Loess and analyzed using proteoDA to perform statistical analysis^89,107^. Linear Models for Microarray Data (limma) with empirical Bayes (eBayes) was used to adjust for the same clinical confounders identified in methylation analysis^89^. Proteins with an FDR-adjusted p-value < 0.05 were considered statistically significant. Pathway enrichment was performed using over-representation analysis using WikiPathways from clusterProfiler^108^. Proteomics pathways with BH-adjusted -log10(q value) > 3 were reported.

### Multi-omic integration of methylation and proteomics data

Convergent genes were identified by intersecting DMPs (limma, BH-adjusted P < 0.05) with differentially abundant proteins (limma, BH-adjusted P < 0.05) with directional concordance (**Supplementary Table 11**). To assess the quantitative relationship between placental DNA methylation and protein abundance, we applied two complementary integration frameworks serving distinct analytical objectives: Sparse CCA and DIABLO integration.

First, to characterise shared axes of co-variation between the two omic layers without imposing subtype labels, we performed Sparse Canonical Correlation Analysis (Sparse CCA) using the *PMA* package^109,110^. The top 5,000 most variable CpGs (ranked by interquartile range) and all quantified proteins were input to a two-block CCA with L1 penalisation applied to both variate weight vectors, with penalty parameters selected by permutation. Three latent variates (K = 3) were extracted, each identifying pairs of methylation and protein linear combinations with maximised cross-block correlation. This unsupervised approach captured general sPE-associated co-regulation between the epigenome and proteome.

Second, to determine whether sPE subtypes defined by differential methylation could be discriminated using a joint epigenetic-proteomic signature, we applied the supervised DIABLO framework in mixOmics package^111^, which simultaneously selects features across omics layers that are maximally correlated within the integration model and discriminative between predefined classes. DIABLO was trained on methylation and proteomic data using sPE subtype labels (sPE_Type1 versus sPE_Type2) as the response, with the number of components and block-specific feature counts tuned by 5-fold cross-validation with 10 repeats. Classification performance was evaluated by balanced error rate (BER). Subtype-specific networks were constructed separately (control vs Subtype 1; control vs Subtype 2; with control as the reference respectively), retaining DIABLO-selected CpG-protein correlations with |r| > 0.4. Genes linked to promoter CpGs and correlated proteins were mapped to STRING v12.0 protein-protein interaction (PPI) networks (STRINGdb v2.14.3; score threshold ≥ 400)^112^, followed by WikiPathways enrichment using enrichR (v3.2)^91^ with BH adjustment. The full WikiPathways list can be found in **Supplementary Table 10**. To reduce pathway redundancy and improve biological interpretability, enriched WikiPathways terms were further aggregated into higher-order functional modules based on shared pathway annotations, overlapping genes and coherent biological functions.

## Data availability statement

Genome-wide DNA methylation data generated in this study, including raw IDAT files and processed methylation matrices, have been submitted to the Gene Expression Omnibus. Proteomics data are available from the corresponding author upon reasonable request. All remaining data supporting the findings of this study are available within the article and its supplementary information files.

## Acknowledgments

We thank the Hawaii Biorepository (HiBR) for providing placental tissue samples, Dr. Yu Liu, a former postdoctoral fellow, for assistance in matching clinical information, the Michigan Institute for Clinical and Health Research (MICHR) for translational research support, and the IDeA National Resource for Quantitative Proteomics at the University of Arkansas for Medical Sciences for proteomics services. This work was supported National Institutes of Health (NIH), National Library of Medicine grants R01 LM012373 and R01 LM012907, the Eunice Kennedy Shriver National Institute of Child Health and Human Development grant R01 HD084633 (to L.X.G.), NIH training grants T32 GM141746 and T32 CA140044 (Advanced Proteogenomics of Cancer) (to Y.D.), and the NIH National Institute of General Medical Sciences grant R24 GM137786 (IDeA National Resource for Quantitative Proteomics). This research was also supported by the National Center for Advancing Translational Sciences of the NIH under Award Number UM1TR004404. The content is solely the responsibility of the authors and does not necessarily represent the official views of the NIH.

## Author contributions

LG envisioned this project, obtained the funding, supervised the study and revised the manuscript. YD designed proteomics assay, performed the data analysis, generated the figures, and wrote the initial manuscript. SL and PB coordinated the multi-omics assays. SL performed sample preprocessing for multi-omics assays. PB, CL, and FA designed the DNA methylation assays. QH provided the deconvolution framework. YL benchmarked HOMED method. JA and JR managed the biorepository and provided the samples. All authors have read the manuscript.

## Reference

1. Karrar, S. A., Martingano, D. J. & Hong, P. L. Preeclampsia. in StatPearls [Internet] (StatPearls Publishing, 2024).

2. Tabassum, S. et al. Preeclampsia and its maternal and perinatal outcomes in pregnant women managed in Bahrain’s tertiary care hospital. Cureus 14, e24637 (2022).

3. Ananth, C. V., Keyes, K. M. & Wapner, R. J. Pre-eclampsia rates in the United States, 1980-2010: age-period-cohort analysis. BMJ 347, f6564 (2013).

4. Magee, L. A. et al. The 2021 International Society for the Study of Hypertension in Pregnancy classification, diagnosis & management recommendations for international practice. Pregnancy Hypertens. 27, 148–169 (2022).

5. Lawless, L., Qin, Y., Xie, L. & Zhang, K. Trophoblast differentiation: Mechanisms and implications for pregnancy complications. Nutrients 15, 3564 (2023).

6. James, J. L., Lissaman, A., Nursalim, Y. N. S. & Chamley, L. W. Modelling human placental villous development: designing cultures that reflect anatomy. Cell. Mol. Life Sci. 79, 384 (2022).

7. Torres-Torres, J. et al. A narrative review on the pathophysiology of preeclampsia. Int. J. Mol. Sci. 25, 7569 (2024).

8. Meng, Y. et al. The role of DNA methylation in placental development and its implications for preeclampsia. Front. Cell Dev. Biol. 12, 1494072 (2024).

9. Youssef, L., Testa, L., Crovetto, F. & Crispi, F. 10. Role of high dimensional technology in preeclampsia (omics in preeclampsia). Best Pract Res Clin Obstet Gynaecol 92, 102427 (2024).

10. Leavey, K., Wilson, S. L., Bainbridge, S. A., Robinson, W. P. & Cox, B. J. Epigenetic regulation of placental gene expression in transcriptional subtypes of preeclampsia. Clinical Epigenetics 10, 28 (2018).

11. Yang, X. et al. Cord blood DNA methylation and cell-type composition are not significantly associated with severe preeclampsia after cell-type and clinical covariate adjustment. Gigascience 15, (2026).

12. Li, B. et al. Identification and validation of new molecular subtypes within the early and late mild cognitive impairment stages of Alzheimer’s disease. medRxiv (2025) doi:10.1101/2023.04.06.23288268.

13. Campbell, K. A. et al. Placental cell type deconvolution reveals that cell proportions drive preeclampsia gene expression differences. Communications Biology 6, 264 (2023).

14. Knihtilä, H. M. et al. Cord blood DNA methylation signatures associated with preeclampsia are enriched for cardiovascular pathways: insights from the VDAART trial. EBioMedicine 98, 104890 (2023).

15. Cirkovic, A. et al. Systematic review supports the role of DNA methylation in the pathophysiology of preeclampsia: a call for analytical and methodological standardization. Biol Sex Differ 11, 36 (2020).

16. Zaki-Dizaji, M., Ebrahimi, A., Saeedinia, R. & Heidary, Z. Epigenetic landscape of placental tissue in preeclampsia: a systematic review of DNA methylation profiles. BMC Pregnancy Childbirth 25, 953 (2025).

17. Layman, C. E. et al. High-throughput methylome analysis reveals differential methylation for early and late onset preeclampsia for mothers and their children. Physiol Genomics 56, 276–282 (2024).

18. Teschendorff, A. E., Breeze, C. E., Zheng, S. C. & Beck, S. A comparison of reference-based algorithms for correcting cell-type heterogeneity in Epigenome-Wide Association Studies. BMC Bioinformatics 18, 105 (2017).

19. Garmire, L. X. et al. Challenges and perspectives in computational deconvolution of genomics data. Nat Methods 21, 391–400 (2024).

20. Jaffe, A. E. & Irizarry, R. A. Accounting for cellular heterogeneity is critical in epigenome-wide association studies. Genome Biol. 15, R31 (2014).

21. Yang, J. I., Kong, T. W., Kim, H. S. & Kim, H. Y. The proteomic analysis of human placenta with pre-eclampsia and normal pregnancy. J. Korean Med. Sci. 30, 770–778 (2015).

22. Sharma, D. D. et al. The management of preeclampsia: A comprehensive review of current practices and future directions. Cureus 16, e51512 (2024).

23. Weissgerber, T. L. & Mudd, L. M. Preeclampsia and diabetes. Curr. Diab. Rep. 15, 9 (2015).

24. Sohlberg, S., Stephansson, O., Cnattingius, S. & Wikström, A.-K. Maternal body mass index, height, and risks of preeclampsia. Am. J. Hypertens. 25, 120–125 (2012).

25. Lindström, L. et al. Chronic hypertension in women after perinatal exposure to preeclampsia, being born small for gestational age or preterm. Paediatr. Perinat. Epidemiol. 31, 89–98 (2017).

26. De Ridder, K., Che, H., Leroy, K. & Thienpont, B. Benchmarking of methods for DNA methylome deconvolution. Nat. Commun. 15, 4134 (2024).

27. Ferro Dos Santos, M. R., Giuili, E., De Koker, A., Everaert, C. & De Preter, K. Computational deconvolution of DNA methylation data from mixed DNA samples. Brief. Bioinform. 25, bbae234 (2024).

28. Houseman, E. A. et al. DNA methylation arrays as surrogate measures of cell mixture distribution. BMC Bioinformatics 13, 86 (2012).

29. Unjitwattana, T., et al. Single-cell RNA-Seq data have prevalent blood contamination but can be rescued by Originator, a computational tool separating single-cell RNA-Seq by genetic and contextual information. bioRxivorg (2025) doi:10.1101/2024.04.04.588144.

30. Teschendorff, A. E., Zhu, T., Breeze, C. E. & Beck, S. EPISCORE: cell type deconvolution of bulk tissue DNA methylomes from single-cell RNA-Seq data. Genome Biology 21, 221 (2020).

31. Liu, Y. scDeconv: an R package to deconvolve bulk DNA methylation data with scRNA-seq data and paired bulk RNA-DNA methylation data. Brief. Bioinform. 23, bbac150 (2022).

32. Liu, Y., Du, Y. & Garmire, L. X. HOMED is Reference-Based DNA Methylation Deconvolution. [Preprint].

33. Kaya, B. et al. Proliferation of trophoblasts and Ki67 expression in preeclampsia. Arch. Gynecol. Obstet. 291, 1041–1046 (2015).

34. Tang, Z. et al. Decreased levels of folate receptor-β and reduced numbers of fetal macrophages (Hofbauer cells) in placentas from pregnancies with severe pre-eclampsia. Am. J. Reprod. Immunol. 70, 104–115 (2013).

35. van Iterson, M., van Zwet, E. W., BIOS Consortium & Heijmans, B. T. Controlling bias and inflation in epigenome- and transcriptome-wide association studies using the empirical null distribution. Genome Biol 18, 19 (2017).

36. Tong, W. & Giussani, D. A. Preeclampsia link to gestational hypoxia. J. Dev. Orig. Health Dis. 10, 322–333 (2019).

37. Yang, X. et al. Gene body methylation can alter gene expression and is a therapeutic target in cancer. Cancer Cell 26, 577–590 (2014).

38. Sanulli, S. et al. Jarid2 Methylation via the PRC2 Complex Regulates H3K27me3 Deposition during Cell Differentiation. Mol Cell 57, 769–783 (2015).

39. Shannon, M. J. et al. Single-cell assessment of primary and stem cell-derived human trophoblast organoids as placenta-modeling platforms. Dev. Cell 59, 776–792.e11 (2024).

40. Charalambous, M. et al. Disruption of the imprinted Grb10 gene leads to disproportionate overgrowth by an Igf2-independent mechanism. Proc. Natl. Acad. Sci. U. S. A. 100, 8292–8297 (2003).

41. Li, Q., Zhu, Y., Yu, X., Shao, X. & Wang, Y.-L. Molecular regulation and functional benefits of trophoblast syncytialization in optimizing maternal-fetal nutrient allocation. Placenta (2025) doi:10.1016/j.placenta.2025.06.003.

42. Tian, G. et al. Expression and function of the LIM homeobox containing genes Lhx3 and Lhx4 in the mouse placenta. Dev Dyn 237, 1517–1525 (2008).

43. Wilson, I., Hamdan, D. D. M., van der Ploeg, R., Perry, T. & Grützner, F. Evolution and expression of glial cells missing (GCM1 and GCM2) in monotremes suggest an ancient role in reproduction. Open Biol 15, (2025).

44. Ushizawa, K. et al. Global gene expression analysis and regulation of the principal genes expressed in bovine placenta in relation to the transcription factor AP-2 family. Reprod Biol Endocrinol 5, 17 (2007).

45. Lynch, C. S. et al. Impact of Placental SLC2A3 Deficiency during the First-Half of Gestation. Int J Mol Sci 23, (2022).

46. Wilson, S. L., Leavey, K., Cox, B. J. & Robinson, W. P. Mining DNA methylation alterations towards a classification of placental pathologies. Hum Mol Genet 27, 135–146 (2018).

47. Wang, T. et al. Epigenome-wide association data implicate fetal/maternal adaptations contributing to clinical outcomes in preeclampsia. Epigenomics 11, 1003–1019 (2019).

48. Anton, L., Brown, A. G., Bartolomei, M. S. & Elovitz, M. A. Differential methylation of genes associated with cell adhesion in preeclamptic placentas. PLoS One 9, e100148 (2014).

49. Martin, E. et al. Epigenetics and Preeclampsia: Defining Functional Epimutations in the Preeclamptic Placenta Related to the TGF-β Pathway. PLoS One 10, e0141294 (2015).

50. Yeung, K. R. et al. DNA methylation profiles in preeclampsia and healthy control placentas. American journal of physiology. Heart and circulatory physiology 310, (2016).

51. Lee, Y. et al. Placental epigenetic clocks: estimating gestational age using placental DNA methylation levels. Aging (Albany NY) 11, 4238–4253 (2019).

52. Mayne, B. T. et al. Accelerated placental aging in early onset preeclampsia pregnancies identified by DNA methylation. Epigenomics 9, 279–289 (2017).

53. Yu, W. et al. Transcriptional regulation of in mouse trophoblast stem cells. Front Cell Dev Biol 10, 918235 (2022).

54. Sharma, R. et al. Temporal control of decidual inflammation by HOXA10 is essential for implantation and its dysregulation is associated with early pregnancy loss. Life Sci 386, 124159 (2026).

55. Sun, L., Dong, Z., Gu, H., Guo, Z. & Yu, Z. TINAGL1 promotes hepatocellular carcinogenesis through the activation of TGF-β signaling-medicated VEGF expression. Cancer Manag Res 11, 767–775 (2019).

56. Eckhart, L., Fischer, H. & Tschachler, E. Phylogenomics of caspase-activated DNA fragmentation factor. Biochem Biophys Res Commun 356, 293–299 (2007).

57. Wenzel, S. E. et al. PEBP1 Wardens Ferroptosis by Enabling Lipoxygenase Generation of Lipid Death Signals. Cell 171, 628–641.e26 (2017).

58. Pijnenborg, R., Vercruysse, L. & Hanssens, M. The uterine spiral arteries in human pregnancy: facts and controversies. Placenta 27, 939–958 (2006).

59. Knöfler, M. & Pollheimer, J. Human placental trophoblast invasion and differentiation: a particular focus on Wnt signaling. Front. Genet. 4, 190 (2013).

60. Ran, J. et al. Hypoxia regulates glycolysis through the HIF-1α/BMAL1/ALDOC axis to reduce oxaliplatin sensitivity in colorectal cancer. J. Cancer 16, 2503–2515 (2025).

61. Li, J. et al. QSOX1 regulates trophoblastic apoptosis in preeclampsia through hydrogen peroxide production. J. Matern. Fetal. Neonatal Med. 32, 3708–3715 (2019).

62. Alfarsi, L. H. et al. SLC1A5 is a key regulator of glutamine metabolism and a prognostic marker for aggressive luminal breast cancer. Sci Rep 15, 2805 (2025).

63. Yuan, V. et al. Cell-specific characterization of the placental methylome. BMC Genomics 22, 6 (2021).

64. Campbell, K. A. et al. Placental and immune cell DNA methylation reference panel for bulk tissue cell composition estimation in epidemiological studies. Epigenetics 19, 2437275 (2024).

65. Lyall, F., Robson, S. C. & Bulmer, J. N. Spiral artery remodeling and trophoblast invasion in preeclampsia and fetal growth restriction: relationship to clinical outcome: Relationship to clinical outcome. Hypertension 62, 1046–1054 (2013).

66. Aouache, R., Biquard, L., Vaiman, D. & Miralles, F. Oxidative stress in preeclampsia and placental diseases. Int. J. Mol. Sci. 19, E1496 (2018).

67. Qi, H., Xiong, L. & Tong, C. Aging of the placenta. Aging (Albany NY) 14, 5294–5295 (2022).

68. Weingrill, R. B. et al. Temporal trends in microplastic accumulation in placentas from pregnancies in Hawai’i. Environ. Int. 180, 108220 (2023).

69. Ho, D. E., Imai, K., King, G. & Stuart, E. A. Matching as Nonparametric Preprocessing for Reducing Model Dependence in Parametric Causal Inference. Political Analysis 15, 199–236 (2007).

70. Nonparametric Preprocessing for Parametric Causal Inference [R package MatchIt version 4.7.2]. (2025).

71. Du, Y. et al. Multiomics analysis of umbilical cord hematopoietic stem cells from a multiethnic cohort of Hawaii reveals the intergenerational effect of maternal prepregnancy obesity and risks for cancers. Gigascience 14, (2025).

72. Aryee, M. J. et al. Minfi: a flexible and comprehensive Bioconductor package for the analysis of Infinium DNA methylation microarrays. Bioinformatics 30, 1363–1369 (2014).

73. Tian, Y. et al. ChAMP: updated methylation analysis pipeline for Illumina BeadChips. Bioinformatics 33, 3982–3984 (2017).

74. Leek, J. T., Johnson, W. E., Parker, H. S., Jaffe, A. E. & Storey, J. D. The sva package for removing batch effects and other unwanted variation in high-throughput experiments. Bioinformatics 28, 882–883 (2012).

75. Pan Du, Richard Bourgon, Gang Feng, Simon Lin. Lumi. (Bioconductor, 2017). doi:10.18129/B9.BIOC.LUMI.

76. Lucas A., Kristina Gervin, Meaghan C., Kelly M., Devin C., John K., Karl T., Robert Lyle, Brock C., Janine Felix. FlowSorted.CordBloodCombined.450k. (Bioconductor, 2019). doi:10.18129/B9.BIOC.FLOWSORTED.CORDBLOODCOMBINED.450K.

77. Hu, Y., Blair, J. D., Yuen, R. K. C., Robinson, W. P. & von Dadelszen, P. Genome-wide DNA methylation identifies trophoblast invasion-related genes: Claudin-4 and Fucosyltransferase IV control mobility via altering matrix metalloproteinase activity. Mol Hum Reprod 21, 452–465 (2015).

78. Fortin, J.-P., Triche, T. J., Jr & Hansen, K. D. Preprocessing, normalization and integration of the Illumina HumanMethylationEPIC array with minfi. Bioinformatics 33, 558–560 (2017).

79. Li, M. et al. DISCO: a database of Deeply Integrated human Single-Cell Omics data. Nucleic Acids Res 50, D596–D602 (2022).

80. Han, X. et al. Construction of a human cell landscape at single-cell level. Nature 581, 303–309 (2020).

81. Liu, Y. et al. Single-cell RNA-seq reveals the diversity of trophoblast subtypes and patterns of differentiation in the human placenta. Cell Res 28, 819–832 (2018).

82. Vento-Tormo, R. et al. Single-cell reconstruction of the early maternal-fetal interface in humans. Nature 563, 347–353 (2018).

83. Cao, J. et al. A human cell atlas of fetal gene expression. Science 370, (2020).

84. Butler, A., Hoffman, P., Smibert, P., Papalexi, E. & Satija, R. Integrating single-cell transcriptomic data across different conditions, technologies, and species. Nat. Biotechnol. 36, 411–420 (2018).

85. Tsoucas, D. et al. Accurate estimation of cell-type composition from gene expression data. Nat Commun 10, 2975 (2019).

86. Dong, M. et al. SCDC: bulk gene expression deconvolution by multiple single-cell RNA sequencing references. Brief Bioinform 22, 416–427 (2021).

87. Salas, L. A. et al. Enhanced cell deconvolution of peripheral blood using DNA methylation for high-resolution immune profiling. Nature Communications 13, 761 (2022).

88. Kazmi, N. et al. Hypertensive Disorders of Pregnancy and DNA Methylation in Newborns. Hypertension (2019) doi:10.1161/HYPERTENSIONAHA.119.12634.

89. Ritchie, M. E. et al. limma powers differential expression analyses for RNA-sequencing and microarray studies. Nucleic Acids Res. 43, e47 (2015).

90. Kasper Daniel Hansen [cre, A. IlluminaHumanMethylationEPICanno.ilm10b4.hg19. (Bioconductor, 2017). doi:10.18129/B9.BIOC.ILLUMINAHUMANMETHYLATIONEPICANNO.ILM10B4.HG19.

91. Xie, Z. et al. Gene Set Knowledge Discovery with Enrichr. Curr Protoc 1, e90 (2021).

92. von Luxburg, U. A Tutorial on Spectral Clustering. (2007).

93. Bolstad, B. M., Irizarry, R. A., Astrand, M. & Speed, T. P. A comparison of normalization methods for high density oligonucleotide array data based on variance and bias. Bioinformatics 19, 185–193 (2003).

94. Kuhn, M. Building Predictive Models inRUsing thecaretPackage. J. Stat. Softw. 28, 1–26 (2008).

95. Friedman, J. H., Hastie, T. & Tibshirani, R. Regularization Paths for Generalized Linear Models via Coordinate Descent. J. Stat. Soft. 33, 1–22 (2010).

96. Prater, M. et al. RNA-Seq reveals changes in human placental metabolism, transport and endocrinology across the first-second trimester transition. Biol. Open 10, bio058222 (2021).

97. Konwar, C. et al. DNA methylation profiling of acute chorioamnionitis-associated placentas and fetal membranes: insights into epigenetic variation in spontaneous preterm births. Epigenetics Chromatin 11, 63 (2018).

98. Mortillo, M., Kennedy, E. G., Hermetz, K. M., Burt, A. A. & Marsit, C. J. Epigenetic landscape of 5-hydroxymethylcytosine and associations with gene expression in placenta. Epigenetics 19, 2326869 (2024).

99. Bhattacharya, A. et al. Placental genomics mediates genetic associations with complex health traits and disease. Nat. Commun. 13, 706 (2022).

100. Shorey-Kendrick, L. E. et al. Impact of vitamin C supplementation on placental DNA methylation changes related to maternal smoking: association with gene expression and respiratory outcomes. Clin. Epigenetics 13, 177 (2021).

101. Wang, W.-J. et al. Genome-wide placental gene methylations in gestational diabetes mellitus, fetal growth and metabolic health biomarkers in cord blood. Front. Endocrinol. (Lausanne) 13, 875180 (2022).

102. Yang, M.-N. et al. Genome-wide placental DNA methylations in fetal overgrowth and associations with leptin, adiponectin and fetal growth factors. Clin. Epigenetics 14, 192 (2022).

103. Essers, R. et al. Prevalence of chromosomal alterations in first-trimester spontaneous pregnancy loss. Nat. Med. 29, 3233–3242 (2023).

104. Khan, A. et al. The application of epiphenotyping approaches to DNA methylation array studies of the human placenta. Res. Sq. (2023) doi:10.21203/rs.3.rs-3069705/v1.

105. Bhatti, G. et al. Placental epigenetic clocks derived from crowdsourcing: Implications for the study of accelerated aging in obstetrics. iScience 28, 113181 (2025).

106. Graw, S. et al. proteiNorm - A User-Friendly Tool for Normalization and Analysis of TMT and Label-Free Protein Quantification. ACS Omega 5, 25625–25633 (2020).

107. Thurman, T. J. et al. proteoDA: a package for quantitative proteomics. J. Open Source Softw. 8, 5184 (2023).

108. Yu, G., Wang, L.-G., Han, Y. & He, Q.-Y. clusterProfiler: an R package for comparing biological themes among gene clusters. OMICS 16, 284–287 (2012).

109. Witten, D. M. & Tibshirani, R. J. Extensions of Sparse Canonical Correlation Analysis with Applications to Genomic Data. Statistical Applications in Genetics and Molecular Biology 8,.

110. Witten, D. M., Tibshirani, R. & Hastie, T. A penalized matrix decomposition, with applications to sparse principal components and canonical correlation analysis. Biostatistics 10, 515–534 (2009).

111. Rohart, F., Gautier, B., Singh, A. & Lê Cao, K.-A. mixOmics: An R package for ’omics feature selection and multiple data integration. PLoS Comput Biol 13, e1005752 (2017).

112. Szklarczyk, D. et al. The STRING database in 2023: protein-protein association networks and functional enrichment analyses for any sequenced genome of interest. Nucleic Acids Res. 51, D638–D646 (2023).

