## Supplementary Figures for "Placental molecular subtypes of severe preeclampsia reveal divergent aging trajectories and fetal growth outcomes"

**Supplementary Figure and Legends**

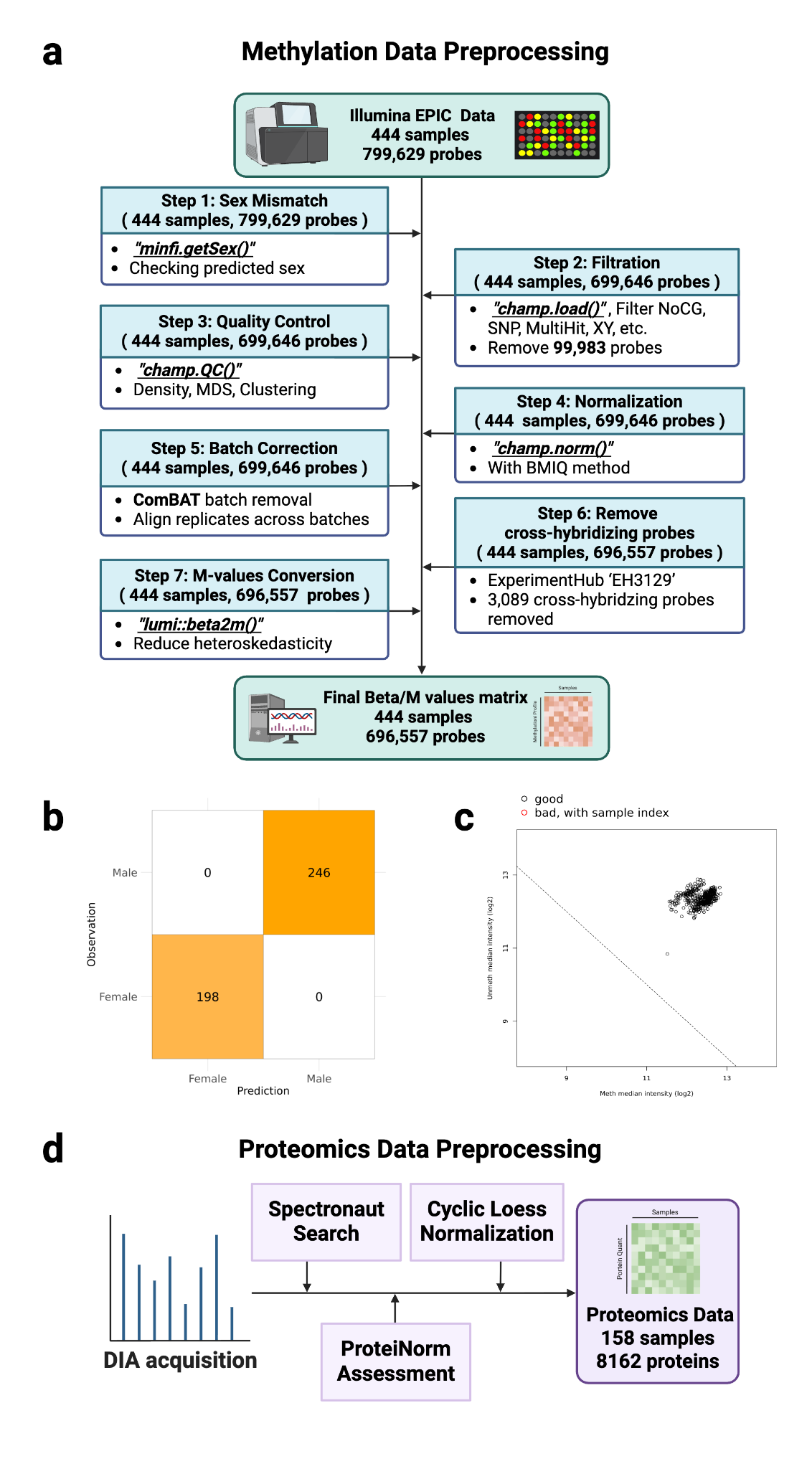

**Supplementary Figure 1. Preprocessing and quality control of placental DNA methylation and proteomics data. (a)** Overview of the DNA methylation preprocessing workflow, including sex-mismatch assessment, probe filtering, sample quality control, BMIQ normalization, ComBat batch correction, removal of cross-hybridizing probes, and conversion to M-values. Created with BioRender. **(b)** Sex-mismatch assessment based on methylation-derived sex prediction. **(c)** Median methylated and unmethylated signal intensity quality control. (d) Overview of the proteomics preprocessing workflow, including DIA-MS acquisition, database annotation, protein-level quality control, and normalization.

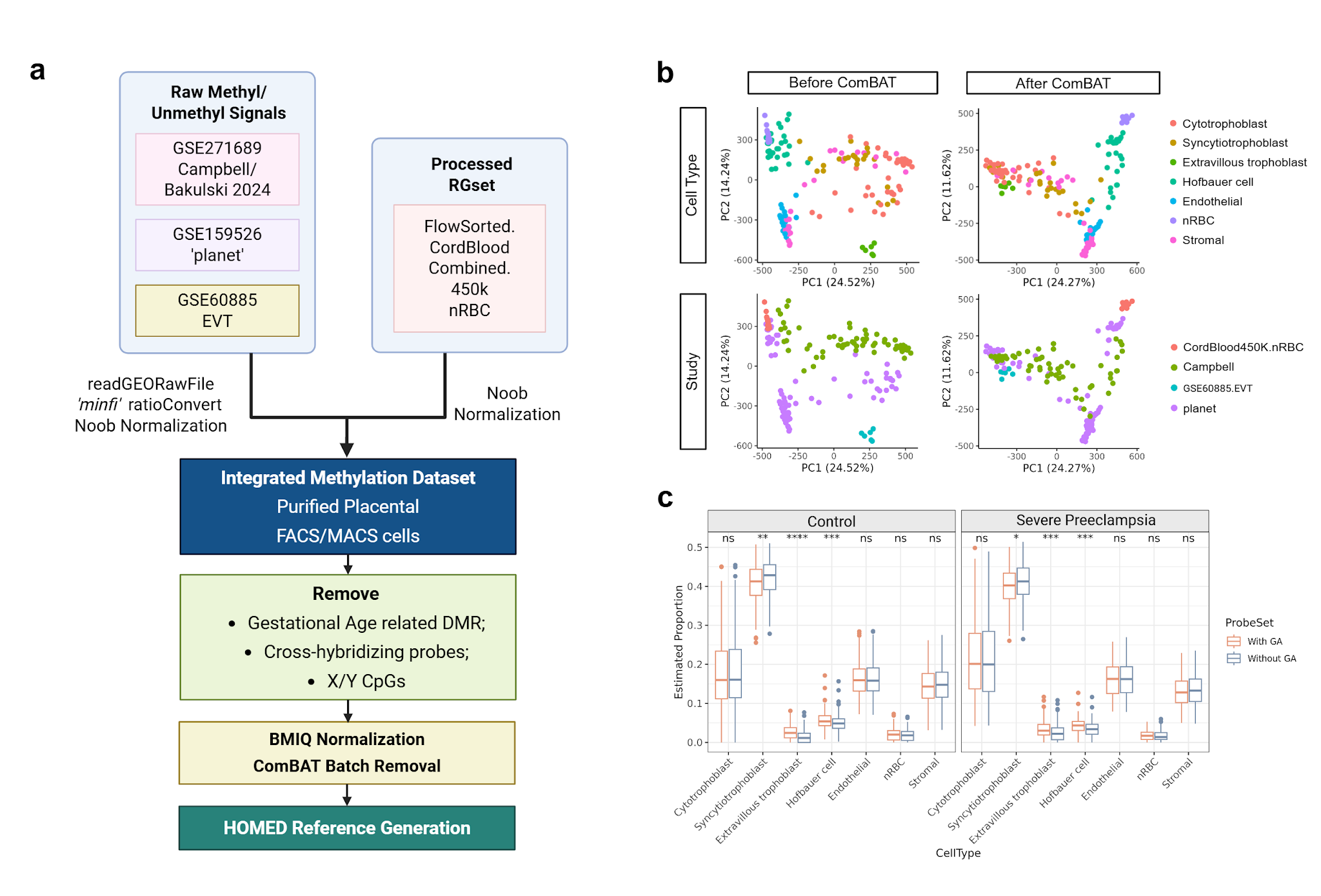

**Supplementary Figure 2. Construction and evaluation of the purified placental methylation reference. (a)** Workflow for integrating purified placental methylation reference datasets used for HOMED placenta reference construction. **(b)** Principal component analysis of purified methylation profiles before and after ComBat correction, colored by cell type and study origin. **(c)** Comparison of HOMED-estimated placental cell-type proportions using methylation references with or without gestational age-associated CpGs. Wilcoxon rank-sum tests were used for group comparisons. *P < 0.05; **P < 0.01; ***P < 0.001; ****P < 0.0001.

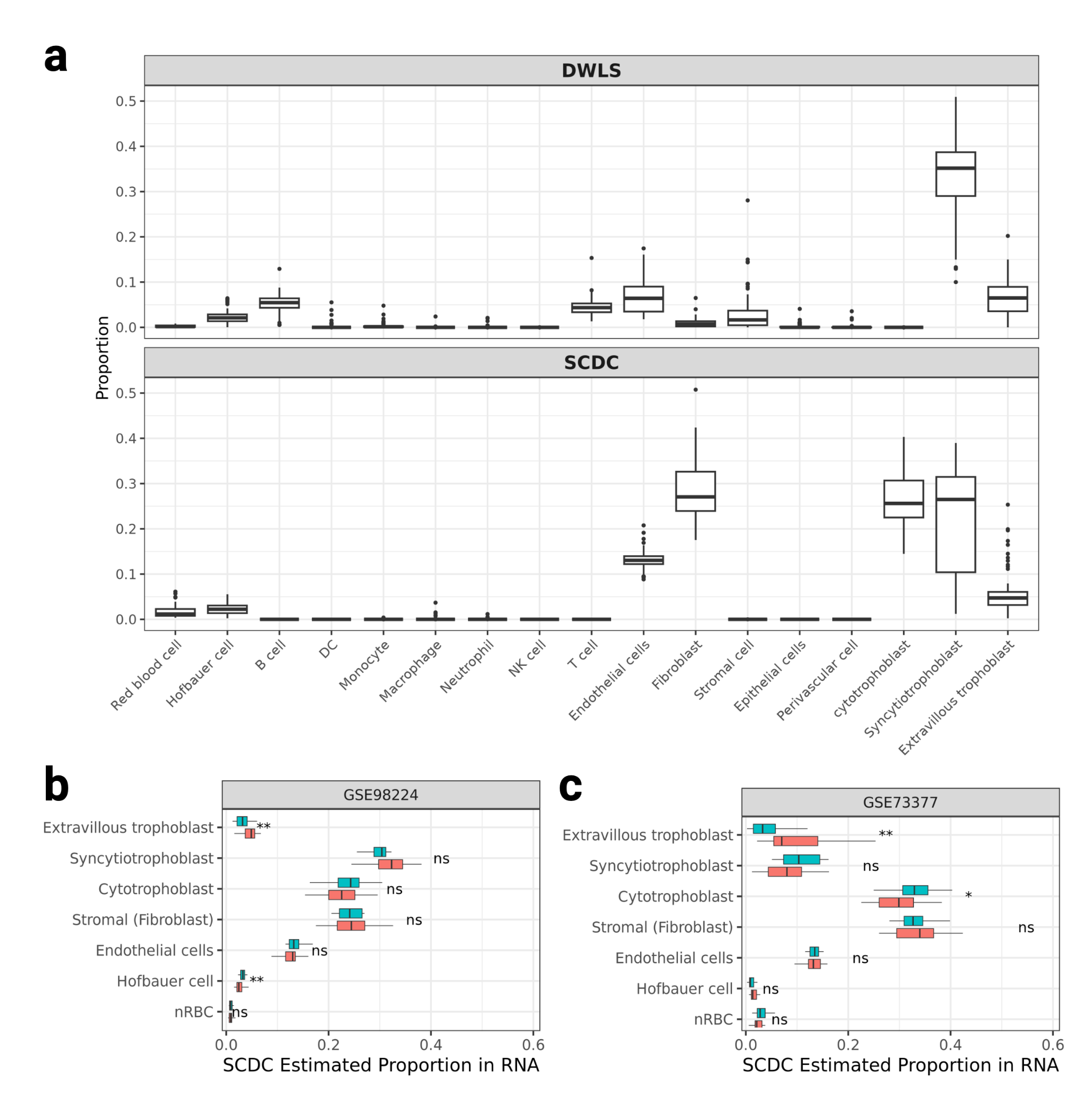

**Supplementary Figure 3. Benchmarking of bulk RNA-seq deconvolution and validation in external PE cohorts. (a)** Comparison of placental cell-type proportion estimates generated by DWLS and SCDC using the integrated placental single-cell RNA-seq reference atlas. **(b,c)** Estimated placental cell-type proportions in two independent PE cohorts, Martin et al. 2015 and Wilson et al. 2018, inferred using SCDC. Wilcoxon rank-sum tests were used for PE versus control comparisons. ns, not significant; *P < 0.05; **P < 0.01; ***P < 0.001; ****P < 0.0001.

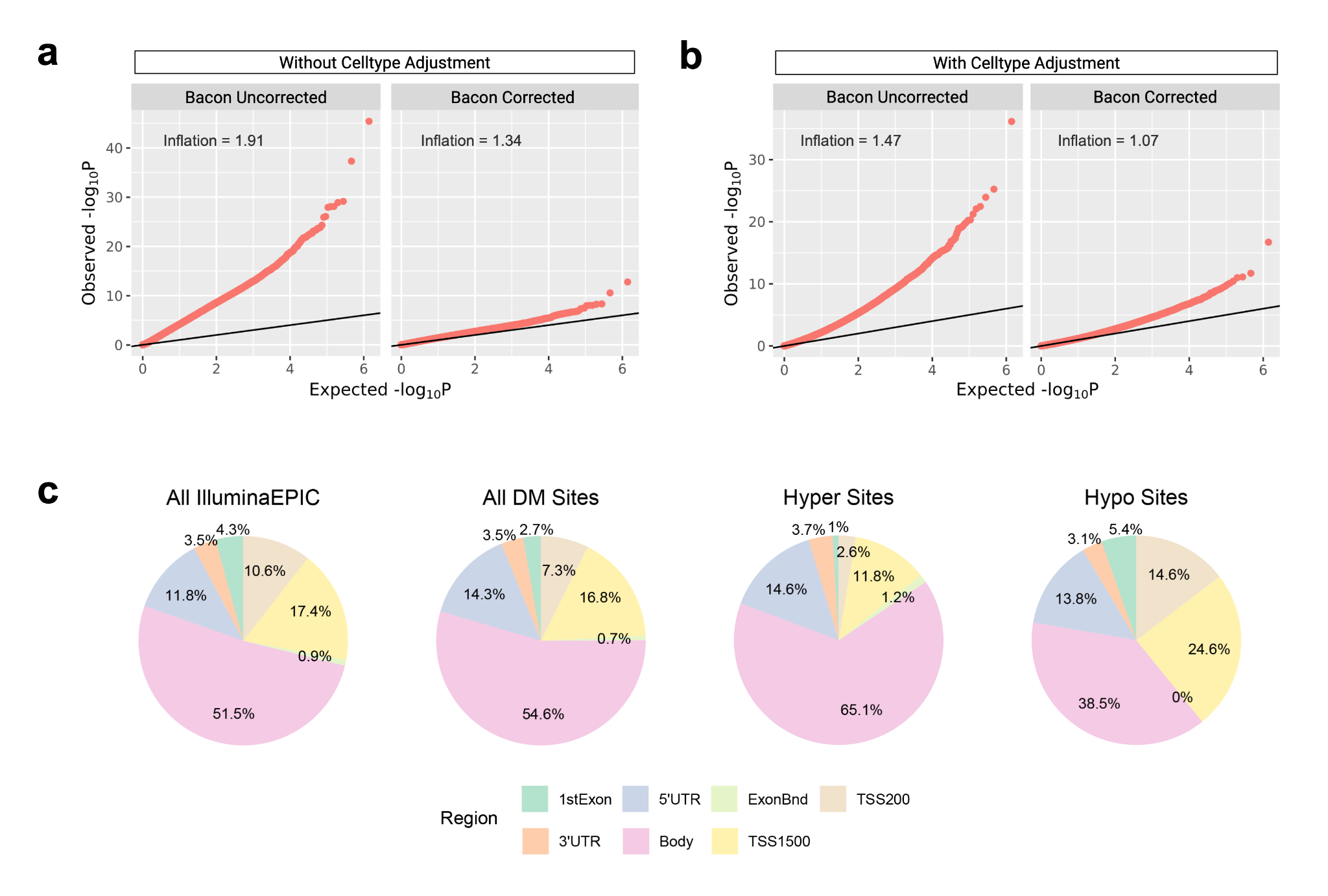

**Supplementary Figure 4. Genomic inflation and genomic annotation of sPE-associated methylation signals. (a)** Quantile–quantile plots of EWAS statistics before HOMED-based cell-type adjustment, shown before and after Bacon correction. **(b)** Quantile–quantile plots of EWAS statistics after HOMED-based cell-type adjustment and Bacon correction. **(c)** Genomic annotation of EPIC array probes and differentially methylated probes, stratified by all significant sites, hypermethylated sites and hypomethylated sites.

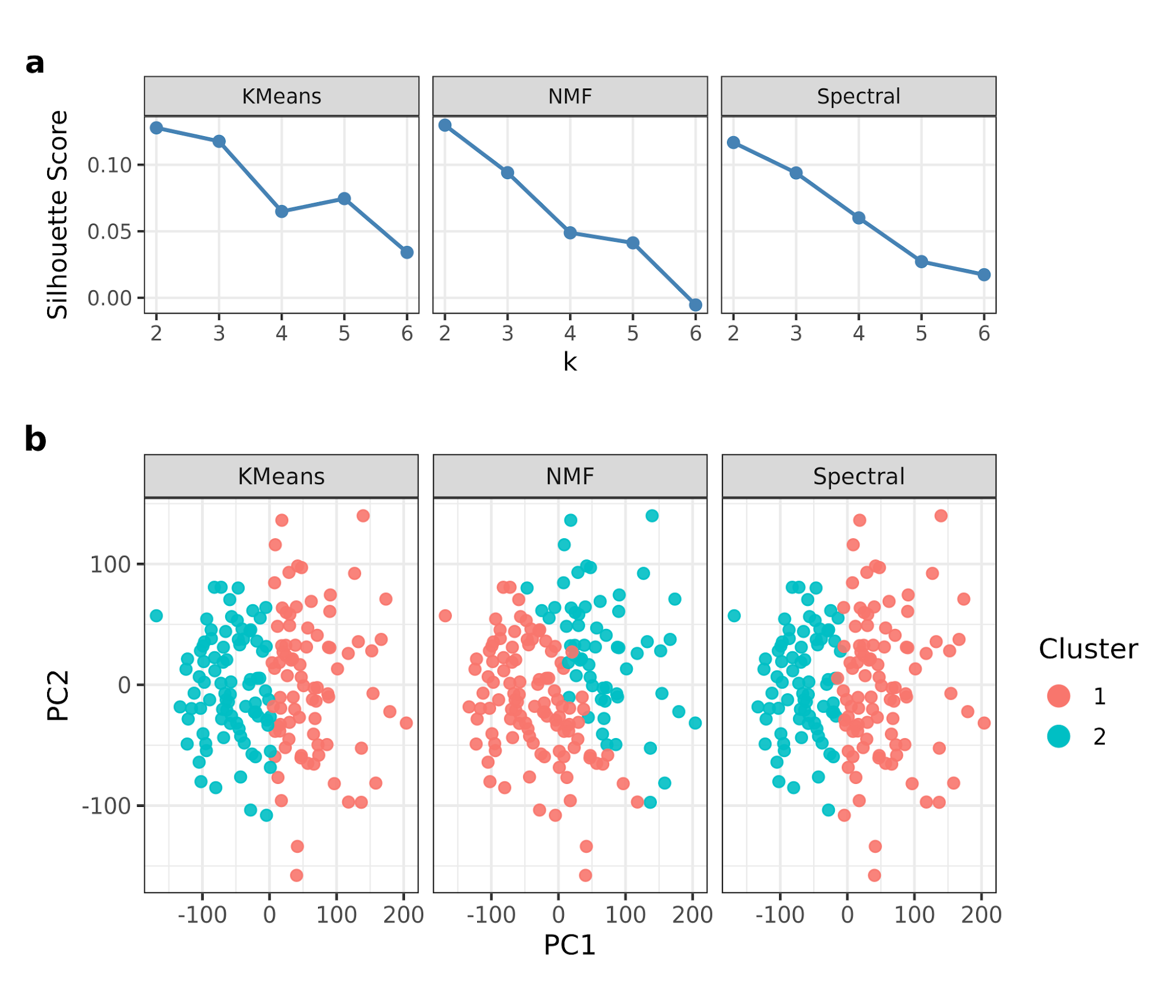

**Supplementary Figure 5. Consensus clustering stability for sPE subtype discovery. (a)** Silhouette scores across clustering methods and candidate cluster numbers used to evaluate subtype stability. **(b)** Principal component analysis of sPE methylation profiles showing subtype assignments from K-means, non-negative matrix factorization and spectral clustering.

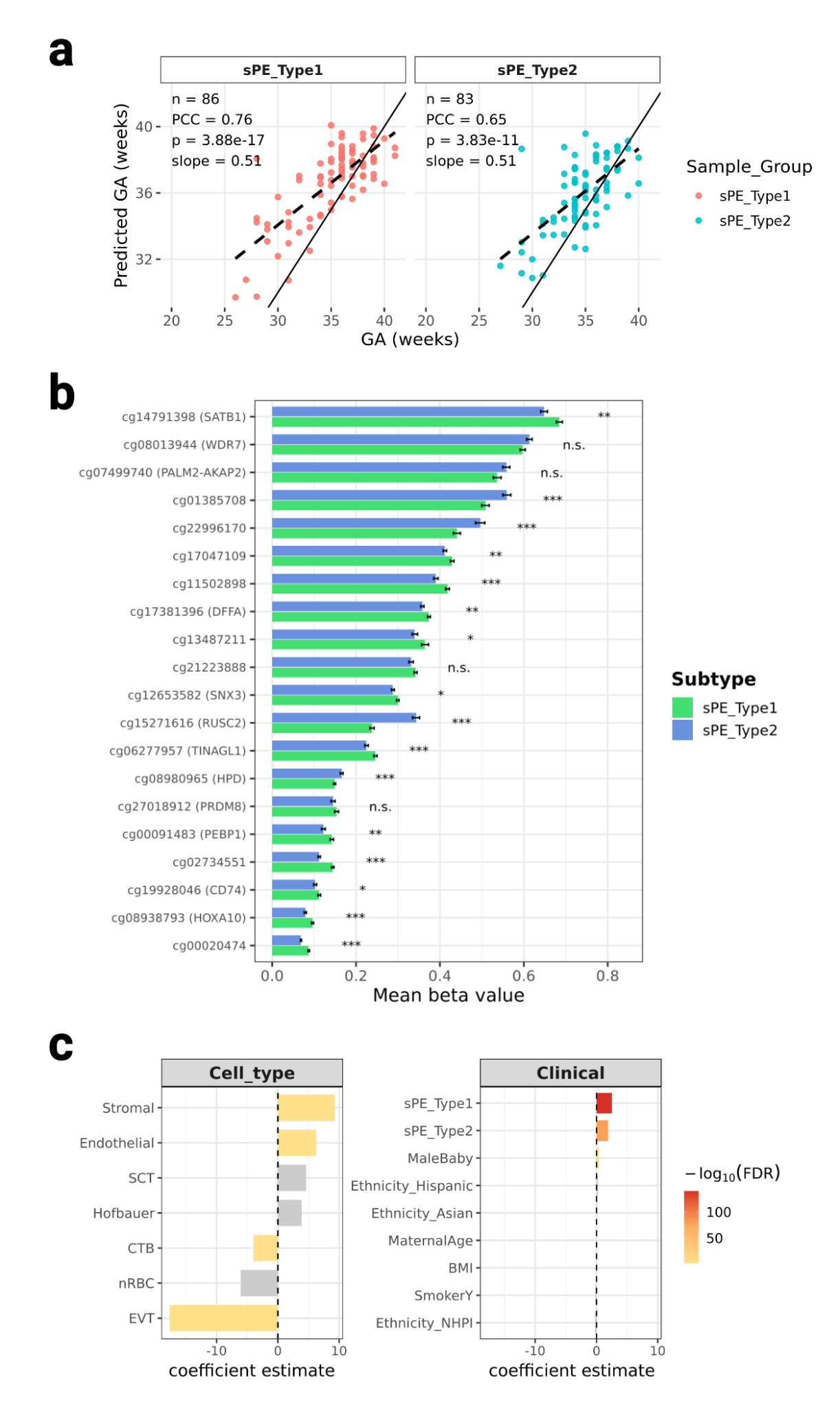

**Supplementary Figure 6. Gestational age clock prediction on sPE subtypes.**Relationship between chronological gestational age and DNAm-predicted biological gestational age in sPE_Type1 and sPE_Type2. Pearson correlation coefficient, P value and regression slope are shown. Dashed lines indicate fitted regression lines; solid lines indicate perfect prediction.

**Supplementary Table Legends**

**Supplementary Table 1. Demographic and clinical characteristics of the methylation cohort.** Clinical and demographic characteristics of propensity-score-matched control and severe preeclampsia placentas used for DNA methylation analysis. Continuous variables are reported as median and interquartile range. P values were calculated using Wilcoxon rank-sum tests for continuous variables and chi-square or Fisher’s exact tests for categorical variables, as appropriate.

**Supplementary Table 2. Placental methylation reference panel summary.** Summary of purified placental methylation reference datasets used for HOMED reference construction, including cell type, study accession, gestational trimester information, and number of collected samples or cells.

**Supplementary Table 3. Top sPE-associated differentially methylated CpGs.** Top 20 hypermethylated and hypomethylated CpG sites associated with sPE after clinical covariate adjustment, HOMED-based cell-type adjustment and Bacon correction. CpG ID, annotated gene symbol, methylation direction, log fold change and Benjamini-Hochberg-adjusted Bacon P value are reported.

**Supplementary Table 4. Clinical and demographic characteristics of sPE methylation subtypes.** Comparison of clinical and demographic variables between sPE Subtype 1 and sPE Subtype 2, including maternal characteristics, pregnancy features, fetal growth outcomes and obstetric complications.

**Supplementary Table 5. Top subtype-associated differentially methylated CpGs.** Top 20 hypermethylated and top 20 hypomethylated CpG sites distinguishing sPE Subtype 2 from sPE Subtype 1. CpG annotations, methylation direction, effect size and adjusted significance values are reported.

**Supplementary Table 6. Elastic-net sPE subtype classifier features.** CpG features included in the elastic-net classifier used to distinguish sPE methylation subtypes. Elastic-net coefficients, genomic annotations and associated gene symbols are reported.

**Supplementary Table 7. Placental gestational age clock dataset summary.** Public and in-house placental methylation datasets used for gestational age clock construction and evaluation. Dataset accession numbers, sample numbers and sample conditions are reported.

**Supplementary Table 8. Elastic-net placental gestational age clock features.** CpG features retained in the elastic-net placental gestational age clock model. Features are reported with annotated gene symbols and model coefficients.

**Supplementary Table 9. Top sparse canonical correlation analysis loadings.** Top 20 DNA methylation and proteomic features contributing to the first two sparse canonical correlation analysis canonical variates. Feature identifiers, annotated genes, omic modality and loading values are reported.

**Supplementary Table 10. PPI-enriched pathways from subtype-specific cross-modal networks.** Protein-protein interaction (PPI)-enriched pathways identified from subtype-specific CpG–protein cross-modal networks. Pathway names, input genes or proteins, enrichment statistics and adjusted significance values are reported.

**Supplementary Table 11. Cross-modal overlapping sPE subtype-associated features.** Gene-level features shared between methylation and proteomic subtype-associated analyses. Direction of DNA methylation change and protein abundance change across sPE subtypes are reported.
